# Chronobiology of anthracycline cardiotoxicity across two biobanks

**DOI:** 10.64898/2026.11.24.26368880

**Authors:** Nadim El Jamal, Thomas G. Brooks, Penn Medicine Biobank, Carsten Skarke, Garret A. FitzGerald

## Abstract

**Background:** Anthracyclines are widely used chemotherapeutics limited by dose-dependent cardiotoxicity. Because cardiovascular physiology and drug handling oscillate across the day, the timing of drug administration may influence cardiac risk. Optimal timing may depend on both an individual’s internal circadian phase and environmental light exposure. We tested whether dosing time is associated with anthracycline cardiotoxicity, whether this is also true of overall treatment outcomes, and whether optimal dosing time is modified by photoperiod and the genetically predicted chronotype.

**Methods:** We analyzed ejection fraction (EF) change following anthracycline exposure in the All of Us research program (AoU; 70 patients) and incident heart failure (HF) in the Penn Medicine Biobank (PMBB; 843 patients). Dosing time was modeled on both clock-time and solar (time-from-sunrise) axes using flexible splines, with likelihood-ratio tests (LRT) for association and for interaction with photoperiod and a germline chronotype polygenic score (cPGS). The cPGS was validated against wearable-derived midsleep and heart-rate rhythms.

**Results:** Dosing time was associated with anthracycline cardiotoxicity in both cohorts, with greater harm at later dosing times (AoU EF LRT p=0.011 clock, p=0.029 sunrise; PMBB HF p=0.052 sunrise, p=0.084 clock). No association was observed with non-HF mortality (p=0.75, 0.77), consistent with a narrowing of the therapeutic index. Photoperiod modified the dosing-time association (AoU p=0.064 clock, p=0.062 sunrise; PMBB p=0.023 clock, p=0.093 sunrise). Modification by cPGS was directionally consistent across cohorts but did not reach significance, likely reflecting attenuation of a polygenic proxy capturing limited chronotype variance.

**Conclusions:** Anthracycline dosing time is associated with cardiotoxicity but not mortality and is modified by photoperiod, identifying dosing time as a modifiable, circadian-sensitive factor. These findings motivate prospective, chronotype- and season-informed studies of anthracycline scheduling.

## Introduction

Anthracyclines are part of therapeutic regimens of multiple types of cancers. However, they are known to cause dose dependent cardiotoxicity.(1) Because multiple aspects of cardiovascular and organismal physiology (including drug targets, metabolizing enzymes, and transporters) oscillate rhythmically throughout the day and night, it has been suggested that drug timing can influence their adverse effect profile.(2, 3) Indeed, in a recent cohort study, patients receiving anthracyclines during the morning hours were at lower risk of developing cancer therapy related cardiac dysfunction compared to patients receiving their doses later in the day.(4) Multiple clocks influence human chrono-physiology: the circadian clock (endogenous molecular clocks), the social clock (local time on a watch), and the solar clock (sunrise-sunset).(5) The solar clock varies through seasons and latitude, and the properties of the circadian clock (such as phase) varies between individuals. Chronotype is the behavioral manifestation of circadian rhythms in sleep/wake time preference.(5) In a GWAS study, 351 loci were associated with chronotype including core molecular clock genes such as *PER1-3*, *CRY* and *ARNTL* with an estimated heritability at ∼ 14%.(6) Seasonal changes affect the duration of melatonin secretion and shift its on- and offset times as well as roughly a quarter of genes in blood and metabolites in serum.(7, 8) Given these three clocks - circadian, social and solar - the question arises whether the optimal dosing time (where oncologic efficacy is maximal and cardiotoxic risk is minimal) is universal or depends on an individual’s internal circadian phase and environmental context. Because clock time does not uniformly reflect internal biological time across individuals or seasons, the cardiotoxic effect of a given dosing hour may vary with chronotype and photoperiod.

In this study, we leverage the convergence of internal (genetic), social, and solar clocks on human cardiovascular physiology to test whether variability in the cardiotoxic effect of anthracycline dosing time can be explained by environmental photoperiod and an individual’s chronotype Across two independent biobanks and using two distinct endpoints, we confirm that dosing time-of-day is associated with cardiotoxicity but not with non-heart-failure mortality. We show that this association is modified by photoperiod and to a lesser extent by genetically predicted chronotype, indicating that the cardioprotective dosing window is not fixed but shifts with an individual’s seasonal light environment.

## Results

### Afternoon dosing increases the risk for cardiotoxicity with no significant effect on survival

From the ejection fraction (EF) data of 70 patients receiving anthracyclines in the AoU research program, we find that dosing time-of-day is significantly associated with post treatment EF change (Likelihood ratio test, LRT p-value = 0.011) with an inverted J-shaped pattern (p-value for non-linearity LRT = 0.043) and significantly drops starting with dosing times around 4pm (Figure 1 A Left). We find similar results when we set our time variable as time from sunrise that changes across months and geographic latitude. (LRT p-value = 0.029, non-linearity p-value = 0.149, Figure 1A Right).

**Figure 1.**
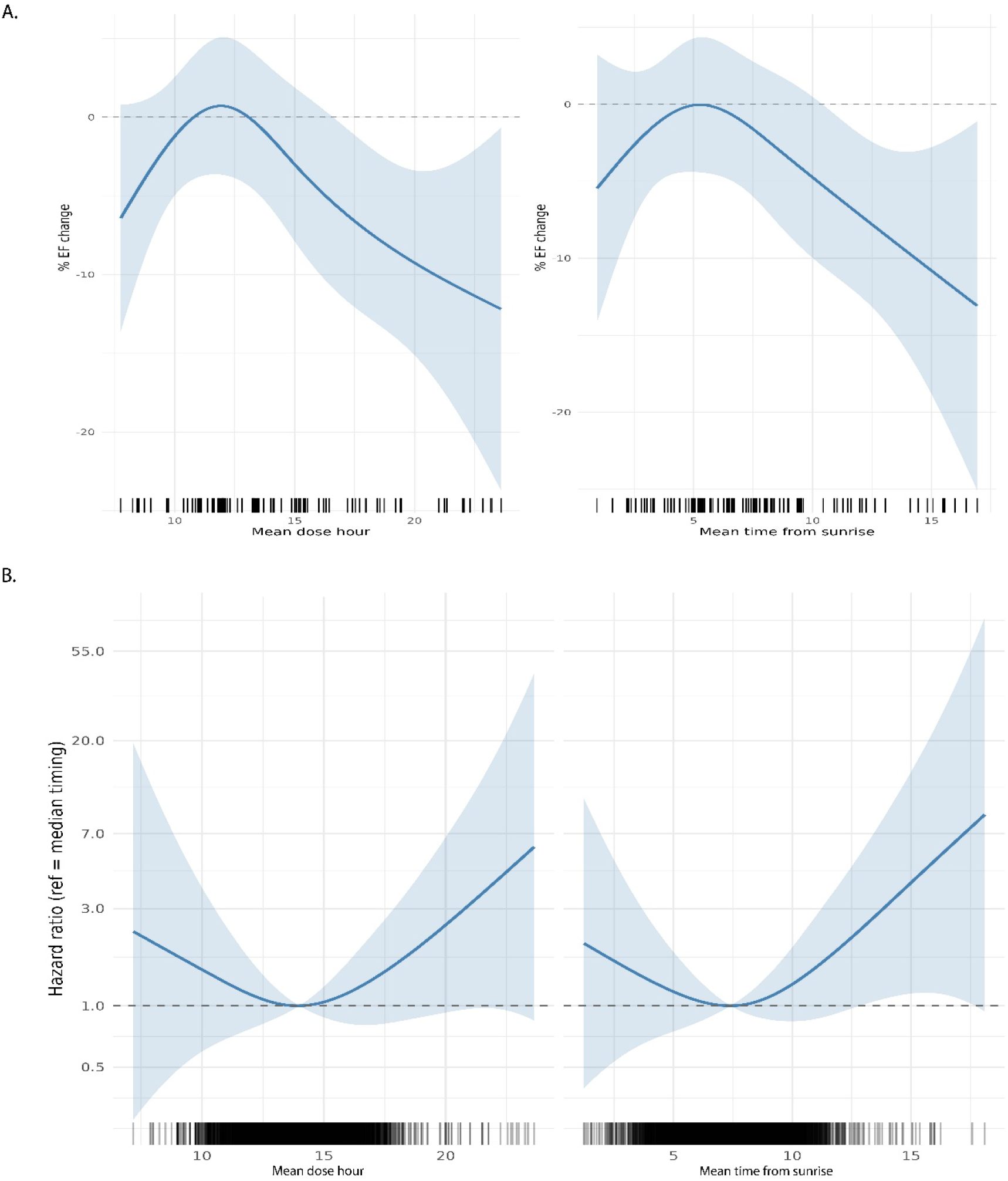
Anthracycline dosing time is associated with cardiotoxicity across two biobanks. (**A**) Predicted percent change in ejection fraction (EF) from baseline as a function of cumulative dose-weighted mean dosing time in 70 anthracycline-exposed AoU participants, modeled with natural splines (3 df). Left, clock time (LRT p = 0.011; non-linearity p = 0.043); right, time from sunrise (LRT p = 0.029; non-linearity p = 0.149). Curves show model fits with 95% confidence intervals (shaded); the dashed line marks no change. (**B**) Hazard ratio for incident heart failure (HF) as a function of dosing time in 843 PMBB patients (47 HF events) from penalized-spline Cox proportional-hazards models, referenced to the median dosing time with 95% confidence intervals. Left, clock time (LRT p = 0.084); right, time from sunrise (LRT p = 0.052; non-linearity p = 0.024). Rugs indicate per-patient dosing-time density. Both outcomes show an inverted-J (EF) / U-shaped (HF) relationship

We then sought to validate the AoU findings on HF diagnosis in an orthogonal Electronic Health Record (EHR) dataset from the Penn Medicine Biobank (PMBB) consisting of 843 patients and 47 HF events (defined as the first appearance of a HF ICD code and a HF specific drug prescription within 7 days) over a median follow up time of 8.52 years. In the independent PMBB HF cohort the significant association seen in AoU was replicated directionally and shows trends mirroring those seen in the AoU EF data with an increase in risk later in the day. The association between clock time and HF risk trends towards significance, (LRT p-value = 0.084), while time from sunrise reaches nominal significance (LRT p-value = 0.052, non-linearity p-value = 0.024, Figure 1B), consistent with lower power for a rarer binary endpoint.

We also examined non-heart failure death to test for a dosing time association with survival. We find no significant association when either dosing parameters were used (LRT p-values = 0.749 for clock time and p=0.767 for time from sunrise, 98 death events, Figure 2).

**Figure 2.**
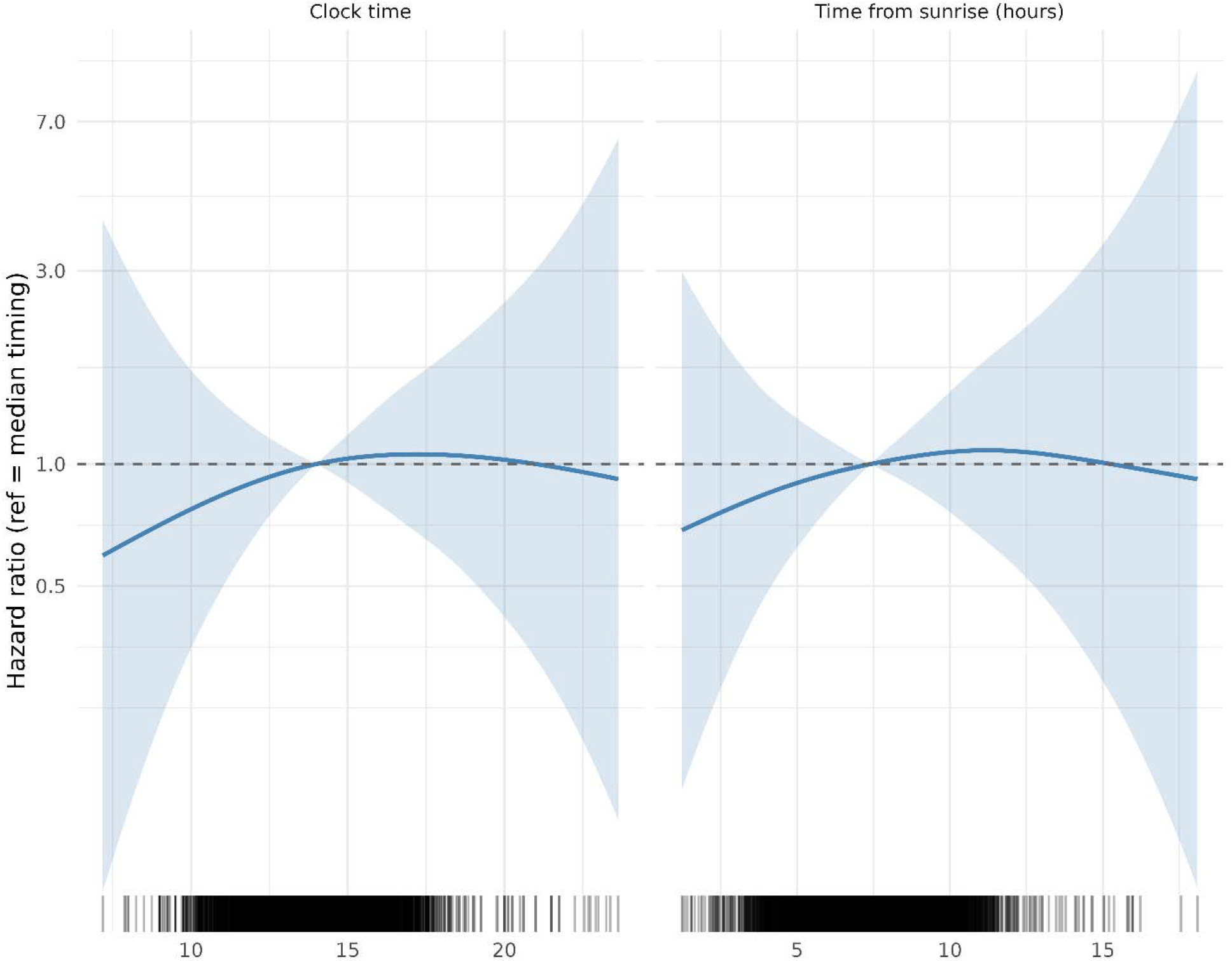
Dosing time is not associated with non-heart-failure mortality. Hazard ratio for non-HF death as a function of dosing time in the PMBB cohort (98 death events), from penalized-spline Cox models referenced to the median dosing time with 95% confidence intervals. Left, clock time (LRT p = 0.749); right, time from sunrise (LRT p = 0.767); rugs show per-patient density. No significant association was observed on either axis.

Thus, we find evidence suggestive of a non-linear association between anthracycline dosing time and cardiotoxicity with no effect on survival. This points to a potential effect on the therapeutic index of anthracyclines. Moreover, because confounding by patient acuity would be expected to track with mortality as well as cardiotoxicity, the absence of any dosing-time association with survival (the more-high powered outcome) argues against such confounding and indicates that our positive cardiotoxicity results are less likely to be biased by patient acuity.

Supplementary figures 1 & 2 show how our sample was selected. Supplementary Tables S1 & S2 describe patient characteristics.

### Photoperiod modifies heart rate rhythms

We sought to test whether the duration of the daily light period (photoperiod: sunset time – sunrise time) modifies the dosing time-of-day association with anthracycline cardiotoxicity. We first validate that photoperiod affects other physiologic rhythms such as heart rate from AoU Fitbit data. We find a significant effect modification for the heart rate cosinor fits by photoperiod length using both clock time and sunrise time anchors (LRT p = <2e-16, Figure 3, Figure S3). Across photoperiod levels from the 5th to 95th percentile (9 to 15 h), the heart-rate rhythm amplitude increased modestly on both axes (clock: 7.58 to 8.05 bpm; sunrise: 7.81 to 8.05 bpm). On the clock axis, the acrophase was essentially stable across photoperiod (14.40 to 14.71 h), whereas on the sunrise axis it shifted markedly later with longer photoperiod, advancing from 7.26 to 9.32h after sunrise. This divergence indicates that the timing of peak heart rate tracks a relatively fixed clock hour rather than a fixed interval after sunrise, consistent with entrainment to clock/social time more than to the solar cycle.(9)

**Figure 3.**
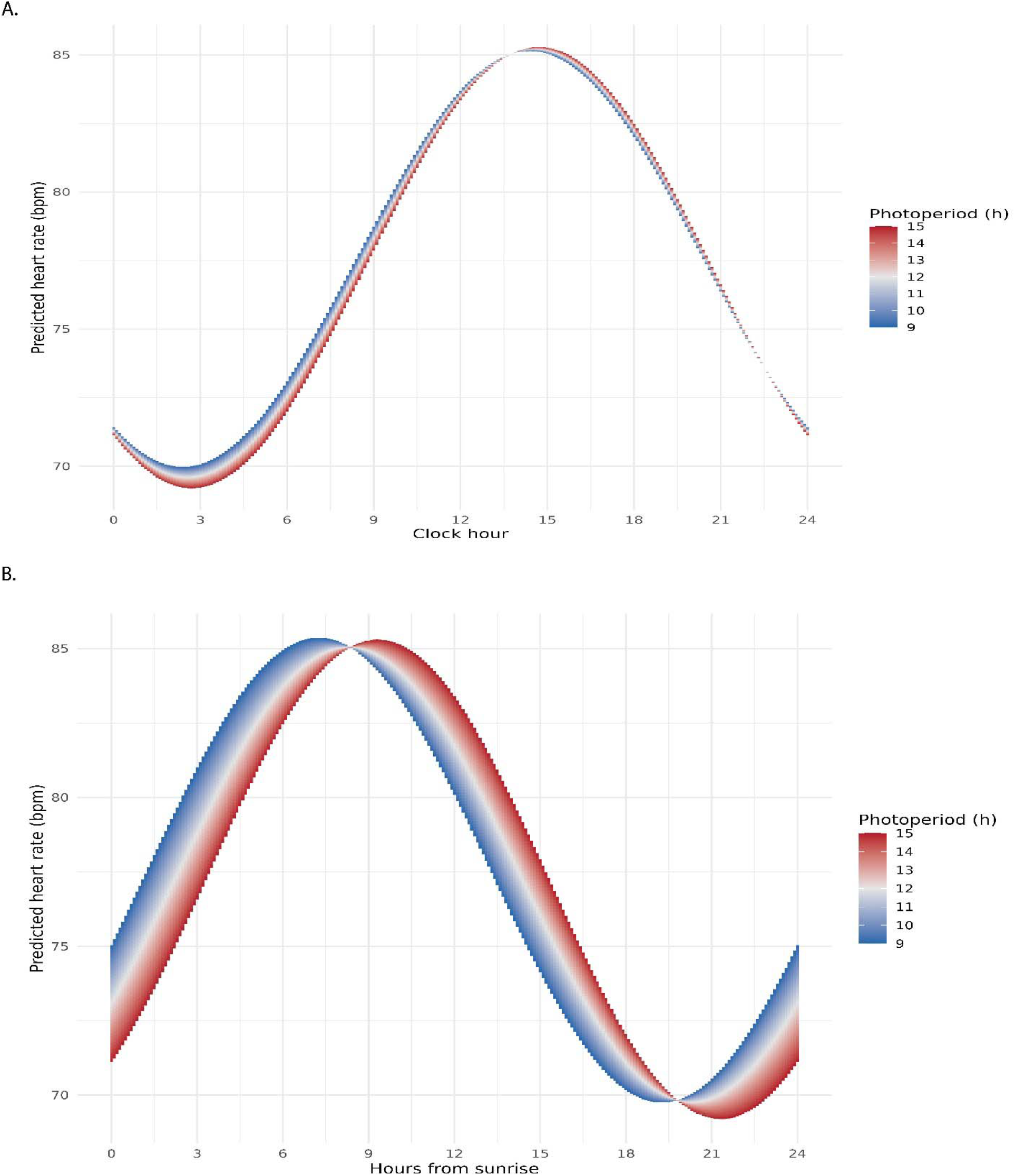
Photoperiod modifies the diurnal heart-rate rhythm. Predicted Fitbit heart rate across photoperiod levels (5th–95th percentile, 9–15 h; color scale) from cosinor mixed-effects models in AoU, on the clock-time (left) and hours-from-sunrise (right) axes; significant effect modification by photoperiod on both axes (LRT p < 2e-16). Longer photoperiod increases rhythm amplitude on both axes; the acrophase is stable in clock time (14.4–14.7 h) but shifts markedly later on the sunrise axis (7.3–9.3 h after sunrise).

### Photoperiod modifies dosing-time–related anthracycline cardiotoxicity

After relaxing the HF definition to only require the ICD code for HF to increase power for interaction tests (843 patients, 133 events), photoperiod also modified the dosing-time association, and within the data-dense interior this modification was directionally concordant across both outcomes. For interaction tests we set the significance level cutoff at 0.1, the threshold followed by Printezi et al.(4) The interaction was significant on both axes in AoU (LRT p = 0.064 clock, p = 0.062 sunrise), significant on the clock axis in PMBB (HF; p = 0.023), and on the solar axis (p = 0.095). Restricting interpretation to the middle of the plot where data are most abundant (as seen in the rugs on Figure 1), the hazard and EF heat bands align well: longer and shorter photoperiods sit on opposite sides of their respective nulls, with a crossover nearmid-afternoon, such that elevated HF hazard corresponds to EF decline and reduced hazard corresponds to EF preservation (Figure 4). The biphasic, double-crossover appearance at the extremes of the heat bands coincides with the sparsely sampled edges. We therefore read photoperiod as a genuine effect modifier. The difference plots are shown over the full observed range (Figure S4). The results from the analysis applying the more stringent HF definition (47 events) show the same directionality but with higher non-significant p-values consistent with power limiting the detection of effect modification in a small event sample (Figure S5).

**Figure 4.**
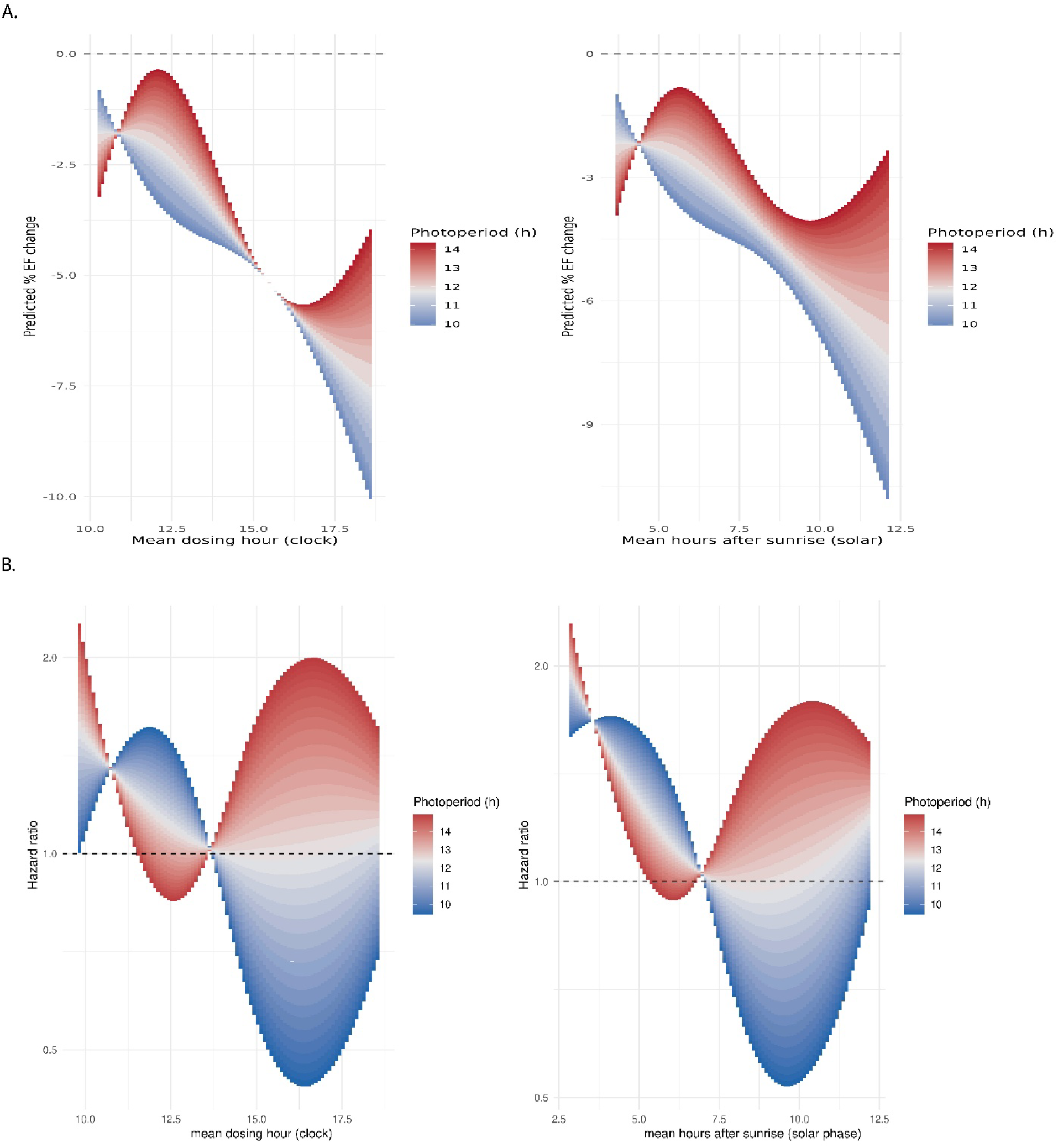
Photoperiod modifies the dosing-time-cardiotoxicity association. Continuous heat bands showing predicted percent EF change (Panel A: AoU; interaction LRT p = 0.064 clock, p = 0.062 sunrise) and HF hazard ratio (Panem B: PMBB; interaction LRT p = 0.023 clock, p = 0.095 sunrise) across dosing time (x-axis) and photoperiod (color scale, 5th–95th percentile), on clock-time and time-from-sunrise axes. Bands are truncated at the 1st–99th percentiles of observed PMBB dosing time (∼15th–85th percentiles in AoU) to limit sparse-data extrapolation; the dashed line marks the null. Within the data-dense interior, longer and shorter photoperiods fall on opposite sides of their respective nulls with a mid-afternoon crossover, concordant across EF and HF. Heat band results in panel B are from Cox proportional hazards models with a less stringent HF definition (843 patients, 133 HF events).

We also tested whether the time-of-day association with non-HF mortality is modified by photoperiod and find no significant effect modification (interaction LRT p = 0.841 clock, p = 0.789 sunrise, figure S6). In addition to shedding light on the therapeutic window, these mortality results also serve as a control against illness acuity.

### Chronotype polygenic score (cPGS) tracks device-measured circadian phenotypes

We next sought to test the effects of chronotype on the dosing time association with cardiotoxicity. Because wearable-derived chronotypes were available in only a small subset of anthracycline-exposed AoU participants who had EF data and were entirely absent from the PMBB validation cohort, we built a chronotype polygenic score as a harmonized, temporally antecedent proxy available in both genotyped cohorts. As a germline instrument, PGS is fixed before exposure and cannot be altered by chemotherapy or cardiac dysfunction, mitigating reverse causation inherent to post-treatment phenotypic chronotype measures.

We built the cPGS using the LDpred2 pipeline from GWAS summary statistics of chronotype in the UK-biobank.(10, 11) Because the UK-biobank participants are mostly of European ancestry while the AoU and PMBB have more diverse participants, we adjusted the cPGS using genetic ancestry principal components (Figure S7, Figure S8). After adjusting the cPGS for genetic ancestry, we tested it on a linearized midsleep time scale (a numeric chronotype measurement defined as the midway point between sleep onset and awakening) from AoU Fitbit data of 62,930 participants. We find that a higher ancestry adjusted cPGS significantly associates with later midsleep times (covariate adjusted p<2e-16, Figure 5A). We also tested the effects of the cPGS on continuous Fitbit heart rate data, another related wearable data stream available in AoU. We find significant effect modification by the cPGS (p<2e-16) when both sunrise and clock times are used as time anchors with a phase delay of about 0.5 hours in the later chronotype participants (peak heart rate at 14.8h, at the 95^th^ percentile cPGS Figures 5B and 5C) compared to earlier chronotype participants, (peak heart rate at 14.3h, at the 5^th^ percentile PGS). We plot the differences between the 95^th^ and 5^th^ PGS percentiles in supplementary Figure S9. We thus validate our polygenic score and establish that participants with later genetically predicted chronotypes have later midsleep times and phase delayed heart rate rhythms compared to participants with earlier chronotypes.

**Figure 5.**
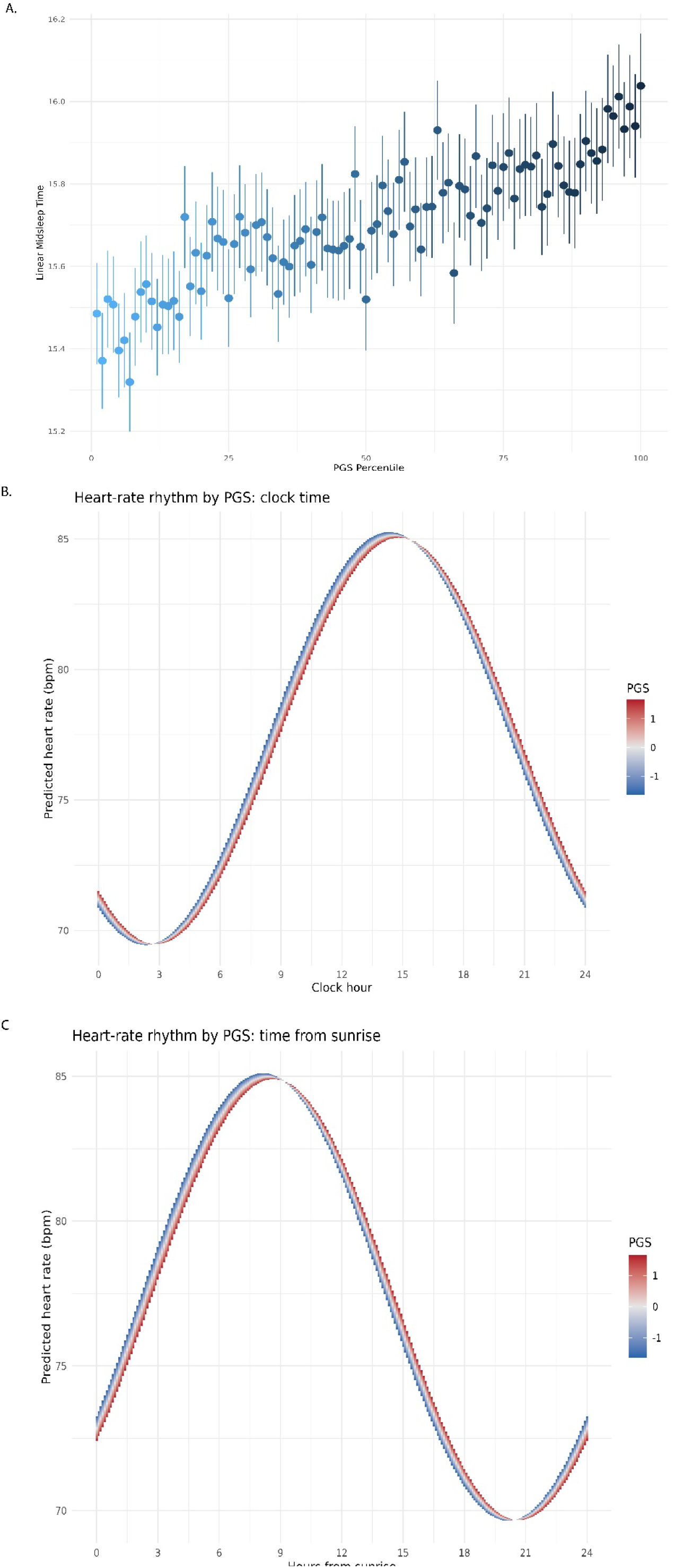
Chronotype polygenic score tracks device-measured sleep and heart-rate rhythms. (**A**) Mean device-derived midsleep time (with 95% confidence intervals) across ancestry-adjusted chronotype PGS percentiles in 47,994 AoU Fitbit participants; higher PGS is associated with later midsleep (covariate-adjusted p < 2e-16). (**B, C**) Predicted Fitbit heart-rate rhythm across PGS levels (5th–95th percentile, color scale) from cosinor mixed-effects models, on the clock-time (**B**) and hours-from-sunrise (**C**) axes; significant effect modification by PGS on both axes (LRT p < 2e-16). Later-chronotype (high-PGS) participants show a ∼0.5 h phase delay in peak heart rate (acrophase 14.8 h at the 95th vs 14.3 h at the 5th percentile).

### Chronotype polygenic score does not significantly modify dosing-time–related anthracycline cardiotoxicity

Across both biobanks, chronotype PGS did not significantly modify the anthracycline dosing time–cardiotoxicity association, though the heat bands were directionally consistent across cohorts. In AoU, the cPGS interaction was not significant on either axis (LRT p = 0.538 clock, p = 0.547 sunrise). The EF heat bands were nonetheless sinusoidal with a single midday crossover: later-chronotype individuals (high PGS) showed greater EF decline with morning dosing and relative preservation with early-afternoon dosing, while earlier chronotypes showed the reverse (Figure 6A). The PMBB HF surfaces mirrored this pattern once the opposing harm conventions are accounted for (elevated HR ≈ EF decline; HR < 1 ≈ EF preservation, Figure 6B), with elevated HF hazard for later chronotypes at morning dosing resolving through the null by mid-afternoon and a single crossover at approximately 14–15 h clock time (≈8–9 h after sunrise), although the PcGS interaction was likewise not significant (p = 0.159 clock, 0.175 solar). This directional convergence of a continuous echocardiographic endpoint and an independent incident ICD-coded endpoint, despite differing populations and confounding structures, is consistent with a "circadian misalignment" interpretation in which dosing discordant with chronotype could be associated with worse cardiac outcomes while aligned dosing appears relatively protective; however, because neither interaction reached significance, we regard this as hypothesis-generating rather than confirmatory. Isolating such an effect will likely require direct phenotypic measurement of chronotype, which would reduce the attenuation inherent to a polygenic proxy that captures only a fraction of chronotype variance. We show difference plots between the 95^th^ and the 5^th^ PGS percentiles in Figure S10.

**Figure 6.**
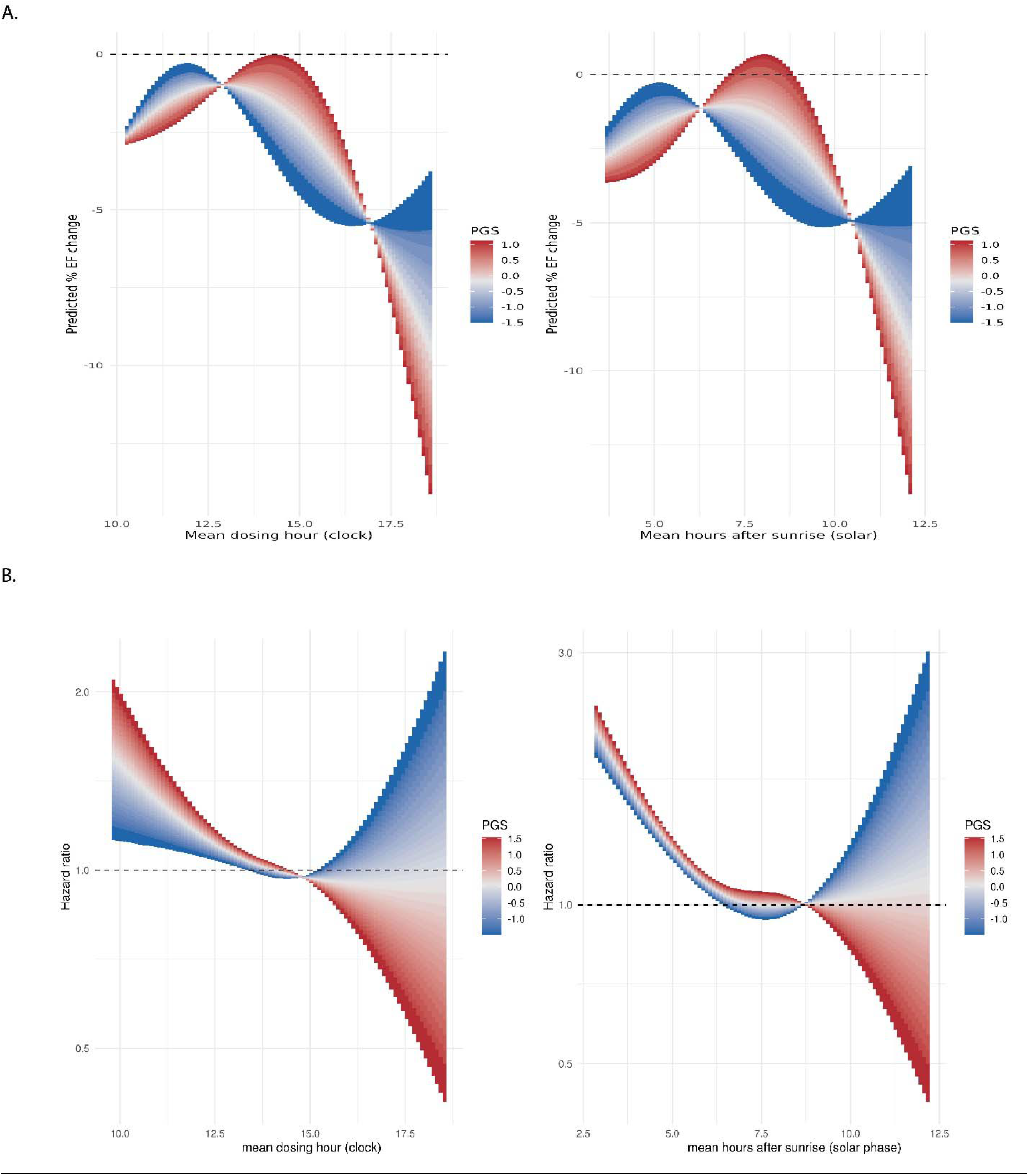
Dosing-time cardiotoxicity stratified by chronotype polygenic score. Continuous heat bands showing the predicted outcome across dosing time (x-axis) and chronotype PGS (color scale, 5th–95th percentile in 2.5% increments).(**A)** predicted percent EF change in AoU (interaction LRT p = 0.547sunrise, p = 0.538 clock); **(B)** HF hazard ratio in PMBB on a log scale (interaction LRT p = 0.159 clock, p = 0.175 sunrise), each referenced to median timing and median PGS. Clock-time and time-from-sunrise axes are shown. Bands are truncated at the 1st–99th percentiles of observed PMBB dosing time (∼15th–85th percentiles in AoU) to limit sparse-data extrapolation; the dashed line marks the null. Later-chronotype individuals show greater harm with morning dosing and relative preservation with early-afternoon dosing; however, interaction tests did not reach significance. Heat band results in panel B are from Cox proportional hazards models with a less stringent HF definition (843 patients, 133 HF events).

Fitting the models applying the stringent HF definitions produces similar results (Figure S11). The chronotype PGS also does not significantly modify the time-of-day association with non-HF mortality (Figure S12).

## Discussion

We show that the cardiotoxic effect of anthracycline dosing time is not fixed but is modified by seasonal light environment and, to a lesser degree, internal circadian phase. Across two independent biobanks and two distinct endpoints, dosing time was associated with cardiotoxicity but not with non–heart-failure mortality. This is consistent with a change in the therapeutic index rather than confounding by patient acuity, and this association was modified by photoperiod and, directionally, by genetically predicted chronotype. These results support the trend reported by Printezi et al. that later time-of-day dosing associates with higher cardiotoxicity risk, (4) and complement their echocardiographic analysis with EHR diagnostic data, ejection fraction changes, and a mortality endpoint. Although animal pharmacokinetic studies (12, 13) led us to hypothesize reduced morning efficacy, we found no association between dosing time and mortality, consistent with recent R-CHOP data showing no significant effect of dosing time on survival after adjustment (14). While overall survival is a commonly reported outcome in oncology efficacy studies, it is a crude measure of efficacy.

Longer photoperiods were associated with larger heart-rate rhythm amplitude, consistent with a more active daytime phase. (15) Peak heart rate remained anchored to a fixed clock hour rather than a fixed interval after sunrise, indicating entrainment predominantly to social/clock time over the solar cycle, in keeping with the influence of socially scheduled activity on cardiovascular rhythms. Photoperiod appeared to modify the dosing-time association more strongly on the clock axis, yet, as with heart rate, it shifted both the magnitude and timing of peak and trough risk. This supports assessing both clock and solar time for insight into the role of diurnal rhythms for parsing benefit-risk ratios.

A later genetically predicted chronotype was associated with a phase delay in the heart-rate rhythm, validating the score against a physiological readout. The chronotype × dosing-time interaction was directionally consistent across both biobanks with later-predicted chronotype patients appearing relatively protected against later dosing, consistent with prior chronotype literature. (16, 17) This association did not reach significance in either cohort. Because chronotype heritability is only ∼14%, (6) a polygenic proxy captures limited variance and likely underestimates the effect; direct phenotypic measurement would provide a more powerful test. Conversely, a germline instrument is fixed before exposure, better approximates a causal effect, and is immune to reverse causation. The consistent directionality across cohorts motivates adequately powered studies using measured chronotype.

A strength of this work is triangulation across independent data types (two biobanks, two cardiotoxicity endpoints, and wearable physiology), such that convergent signals are difficult to attribute to any single cohort. The null mortality association provides a specificity control against confounding by patient acuity. Our exposure-antecedent PGS, built from an independent dataset (UK Biobank) and validated against behavioral (midsleep) and physiological (heart-rate acrophase) readouts, is protected against reverse causation and harmonizable across cohorts lacking wearables. Modeling both clock and solar time guards against wall-clock artifacts.

Limitations include modest, event-limited sample sizes that limit power for the interaction analyses in particular; the PMBB chronotype interaction was directionally concordant with AoU but did not reach significance, though because the polygenic score captures only a fraction of chronotype variance this attenuation is expected to bias interaction estimates toward the null. As an observational study, dosing time was not randomized, and residual confounding from factors associated with both administration timing and outcome cannot be excluded. Exposure and outcomes were derived from real-world data, with anthracycline dose estimated through per-administration counting and doxorubicin-equivalent conversion, and with heart failure and ejection fraction subject to coding, surveillance, and ascertainment biases. Even though we adjust for each of these biases in our models we decrease the risks but do not eliminate them. The polygenic score was derived from a predominantly European-ancestry GWAS, and despite ancestry adjustment its portability across the diverse study populations is imperfect, while geolocation-based estimates of sunrise and photoperiod introduce additional measurement error. Finally, our analyses characterize associations and effect modification but do not causally establish that re-timing administration reduces harm, which will require prospective evaluation.

## Conclusion

Across two independent biobanks and two distinct endpoints, delayed anthracycline dosing was associated with greater cardiotoxicity but not with non-heart-failure mortality, consistent with a narrowing of the therapeutic index rather than confounding by patient acuity. This dosing-time association was modified by photoperiod, while modification by genetically predicted chronotype, though directionally consistent across cohorts, did not reach significance. Together these findings suggest dosing time as a modifiable, circadian-sensitive factor in anthracycline cardiotoxicity and motivate prospective, chronotype- and season-informed studies to test whether optimizing the timing of drug administration can reduce cardiac harm.

## Methods

### Datasets and Ethical Approval

Data for this study was obtained from the All of Us research program (AoU, data version 9) including 535,662 genotyped participants (WGS) and the PennMedicine Biobank (PMBB, data version 4) including 70,493 genotyped participants (genotype array). NE is a credentialed user of the AoU program. AoU research procedures are approved by the AoU IRB and patients provide informed consent before participation. The AoU program determined that use of its data by credentialed users does not constitute human subject research and does not require IRB review. NE applied for and received access to the PMBB data (project ID: 1127). The PMBB is approved by the IRB at the University of Pennsylvania (protocol# 813913) and patients provide informed consent before participation. Our analysis of the PMBB data was deemed exempt from IRB review by the IRB at the University of Pennsylvania (protocol# 859892). For both datasets, only de-identified data was accessed and analyzed.

### Dosing Time and Anthracycline Cardiotoxicity

From both datasets, we selected patients with documented anthracycline exposure of the following drugs: doxorubicin, epirubicin, daunorubicin, idarubicin, and mitoxantrone. We excluded patients who received a liposomal formulation of the drugs, had a non-systemic administration (intra-arterial chemoembolization or intravesical administration), received doses outside 07:00 to 23:59, who had diagnoses where a prolonged infusion may be indicated (Ewing sarcoma, multiple myeloma, Burkitt lymphoma, osteosarcoma, soft tissue sarcoma, liposarcoma, leiomyosarcoma, synovial sarcoma, fibrosarcoma, angiosarcoma, rhabdomyosarcoma, myxofibrosarcoma, pleomorphic sarcoma, diffuse large b-cell lymphoma, primary mediastinal lymphoma), and where we could not estimate the anthracycline dose. These exclusions decrease the risk of confounding by indication. The ability to estimate the anthracycline dose increases confidence that a patient received the dose at the associated timestamp.

Anthracycline dose was estimated by counting each recorded administration day as one dose of the regimen’s published adult per-dose amount (mg/m², lower bound if a range)(18) and converting other anthracyclines to doxorubicin-equivalents (epirubicin 0.8, daunorubicin 0.6, idarubicin 4.5, mitoxantrone 10.5).(4) Regimens were identified from anti-neoplastic partner drugs given within 2 days of the anthracycline (with dosing intervals used only to distinguish identical drug combinations), and unresolved administrations were imputed to match a patient’s other administrations. Including cumulative dose in our models thus helps adjust for the cancer type and treatment regimen.

Dosing time was either determined as the timestamp reported in the EHR for drug administration or as time from sunrise. Sunrise time was calculated using the R package ‘Suncalc’ based on date and longitude and latitude. For the PMBB that has data from a Philadelphia based health system, longitude and latitude were fixed at -75.1652 and 39.9526. Because AoU contains data from patients across the United States each person’s longitude and latitude was estimated from population weighted Zip code centroids. A cumulative mean time weighted by total cumulative dose contribution was then calculated for each dose.

In PMBB outcomes were defined as the earliest occurrence of an ICD code for heart failure (I42.7, I43, I42.9, I50.4, I42.0, I50.9, I42.8, I50.8, I50.2) that is accompanied by a HF specific drug prescription within 7 days. HF-specific drugs included: sacubitril/valsartan, mineralocorticoid receptor antagonists (spironolactone, eplerenone, finerenone); epithelial sodium channel (ENaC) blockers (amiloride, triamterene); ivabradine; vericiguat; digoxin; dobutamine; milrinone; loop diuretics (furosemide, bumetanide, torsemide, ethacrynic acid); and thiazide and thiazide-like diuretics (chlorothiazide, metolazone). The addition of HF drugs to the definition of HF increases the specificity of our outcome assessment. Patients with HF codes prior to their first dose were excluded. We then fitted Cox proportional hazards models with time varying covariates with each period in follow up starting at the chemotherapy cycle time and ending at either the next administration or at censoring. The dosing time variables were included as penalized splines. The models were adjusted for cumulative anthracycline dose, an indicator for daylight savings time, age, sex, the number of days with EHR encounters (controlling for detection bias), and the first 3 genetic ancestry principal components. To avoid overfitting with a small event number, we limited the covariate list to the above. We censored at the date of the last EHR encounter on record, death, or HF incidence. The proportional hazards assumption was tested using Shoenfeld residuals. To test whether the dosing time variable significantly associated with HF risk, we ran likelihood ratio tests comparing the fully adjusted models with and without the time variables. We then plotted the model fits with 95% confidence intervals using the coefficients and the covariance matrices.

From the AoU dataset, we used ejection fraction (EF) values reported from cardiac imaging modalities. We excluded patients with a minimum EF lower than 50% for the period before the first dose. We divided the follow-up time into periods starting with each chemotherapy cycle and ending with either the next cycle or, for the final cycle, the end of available echocardiographic follow-up, and assigned each echocardiogram performed after a cycle’s administration (up to and including the next administration day) to that cycle, retaining the lowest ejection fraction per period. Periods without an echocardiogram were excluded from the analysis, but their doses were still counted toward the cumulative dose. We then fit mixed effects regression models with the percent change in EF from baseline as the dependent variable and the natural spline terms of the dosing time variables with 3 degrees of freedom as the independent variable of interest. Covariates included in the models were: cumulative dose, sex, age, baseline EF, the time of the nadir EF from the first dose, the number of EF measurements in the period, a daylight savings time indicator, and the first 2 genetic ancestry principal components. Similar to the PMBB analysis above we tested the significance of the dosing time association using likelihood ratio tests and plotted the model fits with 95% confidence intervals.

### Building and Testing the Chronotype Polygenic Score

We obtained publicly available summary statistics from a GWAS for chronotype (phenotype 1180 in the UK biobank with levels of: Definitely a morning person, more morning than evening, more evening than morning, and definitely an evening person).(19) The GWAS was run on 301,143 participants, included 1,0894,596 variants, and was adjusted for age, sex, and the first 20 ancestry principal components.(10) From these statistics we used LDpred2-auto to compute polygenic score coefficients that account for linkage disequilibrium based on the HapMap3 plus reference panel.(11) This resulted in 1,385,528 variants contributing to our polygenic score (PGS).

Based on the availability of genetic data, we used whole genome sequencing data in AoU and imputed array data in the PMBB (1,354,610 matched variants). We included variants with a genotype missingness rate of < 10% and a minor allele frequency ≥1% thus retaining1,261,731 variants in Aou and 1,246,331 variants in the PMBB. For individuals with missing variant genotypes we used applied mean imputation. We use the BigProdVec function from the bigsnpr R package to multiply each PGS coefficient with its corresponding variant genotype. The PGS was adjusted for genetic ancestry. Within each dataset, we regressed the PGS on the first 16 principal components (PCs), then modeled the squared residuals on the same PCs using a gamma generalized linear model with a log link to keep predicted variances positive. The adjusted PGS was each residual divided by the square root of its predicted variance, giving an approximately mean-zero, unit-variance score across ancestry backgrounds.(20)

We then tested the adjusted PGS using sleep Fitbit data available in the AoU program from 62,930 participants. Of these we selected genotyped participants with at least 8 weekend days of data and who did not have sleep onset times between 06:00 and 18:00 or sleep durations <3H or >15h thus retaining 47,994 participants. We used the circular package in R to calculate each person’s mean midsleep time (midpoint between sleep onset and awakening) and then transformed the circular midsleep to a linear variable with increasing midsleep time by subtracting 12 from midsleep times at night before midnight and adding 12 to those after. We then plotted the adjusted PGS percentiles against the mean and confidence interval of the midsleep of each percentile and fit a linear model for midsleep adjusted for age, sex, and the first 10 PCs.

### Effect Modification by Chronotype PGS and Photoperiod

We calculated photoperiod by subtracting sunrise time from sunset time obtained from the suncalc package. For each timing axis (clock time and solar phase, i.e. hours after sunrise) and each candidate modifier (chronotype PGS and photoperiod), we fit a model with the natural-spline timing term (3 df) entered additively with the modifier and a model with a full timing-spline × modifier interaction and compared the two by likelihood-ratio test (3 df) to assess effect modification. For interaction tests we use a p-value cutoff of 0.1 because interaction tests are usually underpowered.(4) We used the same model structures we used to test for the time-of-day association with cardiotoxicity above. Because interaction tests are usually underpowered, we expanded the definition of heart failure in the PMBB to only include the presence of an ICD code for HF. We adjusted for the same covariates as above except for including only 1 PC for models testing modification by cPGS and no PCs for models testing photoperiod modification.

We visualized the fitted interaction as a continuous heat band by predicting the outcome (log hazard ratio in PMBB, predicted % EF change in AoU) over a grid of dosing times crossed with the modifier evaluated from its 5th to 95th percentile in 2.5% increments, holding covariates at reference values (medians for continuous covariates, the modal level for categorical ones), and shading each timing × modifier cell by the modifier value. PMBB predictions are expressed as hazard ratios relative to a single common reference (median timing and median modifier) on a log scale, and the band was drawn over the 1st–99th percentiles of observed dosing time so that sparsely sampled tails did not obscure the interior pattern. This corresponded to 15^th^ to 85^th^ percentiles of observed dosing time in AoU.

To summarize the strength of modification across dosing time, we contrasted the high-versus low-modifier prediction (95th vs. 5th percentile) at each timing value and plotted this difference with a pointwise 95% confidence interval. The contrast and its variance were obtained from the difference in model design matrices multiplied by the coefficient vector and its covariance matrix, yielding the log-HR difference (exponentiated to a hazard ratio) in PMBB and the difference in predicted % EF change in AoU; these plots were drawn over the full observed range of dosing times, with a rug showing the per-patient data density.

### Heart Rate Rhythm Modification by Chronotype PGS and Photoperiod

We extracted minute level Fitbit heart rate data from AoU participants with at least 7 days of monitoring (61,693 participants). We averaged all hourly measurements for every person-date-hour combination. We calculated sunrise and photoperiod in similar ways as above using the suncalc package.

We modeled person × time-bin mean heart rate (weighted by the number of contributing observations) using linear mixed-effects models (lmer) with a cosinor parameterization, representing time-of-day by paired 24-hour cosine and sine terms on two axes: clock hour and hours from sunrise. Each model included a per-person random intercept and was adjusted for sex, birth year, and the first ten genetic principal components; and a modifier (chronotype PGS, photoperiod) interacted with both cosine and sine terms. Effect modification of the rhythm was tested by likelihood-ratio test (2 df) comparing an additive model to the cosinor × modifier interaction, both fit by maximum likelihood. For visualization, model-predicted heart rate (random effects set to zero) was computed over a grid of times of day crossed with the modifier from its 5th–95th percentile, holding covariates at reference values, and rendered as a heat band shaded by modifier value with a shared color scale and y-axis across panels. From the interaction coefficients we derived rhythm amplitude (√(β_cos² + β_sin²)) and acrophase (atan2(β_sin, β_cos), rescaled to hours) at the 5th, 50th, and 95th modifier percentiles.

### Data Availability

All summary statistics and code used are available upon reasonable request to the authors. Patient level data cannot be shared. Readers are directed to the All of Us Research Program and the Penn Medicine Biobank to apply for and access patient level data.

### Funding and Conflict of Interest

This work did not receive any funding. Funding sources for the two biobanks are listed below in the Acknowledgments. The authors report no competing financial interests.

## Acknowledgements

The All of Us Research Program is supported by the National Institutes of Health, Office of the Director: Regional Medical Centers: 1 OT2 OD026549; 1 OT2 OD026554; 1 OT2 OD026557; 1 OT2 OD026556; 1 OT2 OD026550; 1 OT2 OD 026552; 1 OT2 OD026553; 1 OT2 OD026548; 1 OT2 OD026551; 1 OT2 OD026555; IAA #: AOD 16037; Federally Qualified Health Centers: HHSN 263201600085U; Data and Research Center: 5 U2C OD023196; Biobank: 1 U24 OD023121; The Participant Center: U24 OD023176; Participant Technology Systems Center: 1 U24 OD023163; Communications and Engagement: 3 OT2 OD023205; 3 OT2 OD023206; and Community Partners: 1 OT2 OD025277; 3 OT2 OD025315; 1 OT2 OD025337; 1 OT2 OD025276. In addition, the All of Us Research Program would not be possible without the partnership of its participants.

We acknowledge the Penn Medicine BioBank (PMBB) for providing data and thank the patient-participants of Penn Medicine who consented to participate in this research program. We would also like to thank the Penn Medicine BioBank team and Regeneron Genetics Center for providing genetic variant data for analysis. The PMBB is approved under IRB protocol# 813913 and supported by Perelman School of Medicine at University of Pennsylvania, a gift from the Smilow family, and the National Center for Advancing Translational Sciences of the National Institutes of Health under CTSA award number UL1TR001878.

## Penn Medicine BioBank Banner Author List and Contribution Statements

### PMBB Leadership Team

Daniel J. Rader, M.D., Marylyn D. Ritchie, Ph.D.

## Contribution

All authors contributed to securing funding, study design and oversight. All authors reviewed the final version of the manuscript.

### Patient Recruitment and Regulatory Oversight

JoEllen Weaver, Nawar Naseer, Ph.D., M.P.H., Giorgio Sirugo, M.D., P.h.D., Afiya Poindexter, Yi-An Ko, Ph.D., Kyle P. Nerz

## Contributions

JW manages patient recruitment and regulatory oversight of study. NN manages participant engagement, assists with regulatory oversight, and researcher access. GS assists with researcher access. AP, YK, KPN perform recruitment and enrollment of study participants.

### Lab Operations

JoEllen Weaver, Meghan Livingstone, Fred Vadivieso, Stephanie DerOhannessian, Teo Tran, Julia Stephanowski, Salma Santos, Ned Haubein, P.h.D., Joseph Dunn

## Contribution

JW, ML, FV, SD conduct oversight of lab operations. ML, FV, AK, SD, TT, JS, SS perform sample processing. NH, JD are responsible for sample tracking and the laboratory information management system.

### Clinical Informatics

Anurag Verma, Ph.D., Colleen Morse Kripke, M.S. DPT, MSA, Marjorie Risman, M.S., Renae Judy, B.S., Colin Wollack, M.S.

## Contribution

All authors contributed to the development and validation of clinical phenotypes used to identify study subjects and (when applicable) controls.

### Genome Informatics

Anurag Verma Ph.D., Shefali S. Verma, Ph.D., Scott Damrauer, M.D., Yuki Bradford, M.S., Scott Dudek, M.S., Theodore Drivas, M.D., Ph.D.,

## Contribution

AV, SSV, and SD are responsible for the analysis, design, and infrastructure needed to quality control genotype and exome data. YB performs the analysis. TD and AV provides variant and gene annotations and their functional interpretation of variants.

## Supplementary Figures

**Figure S1.**
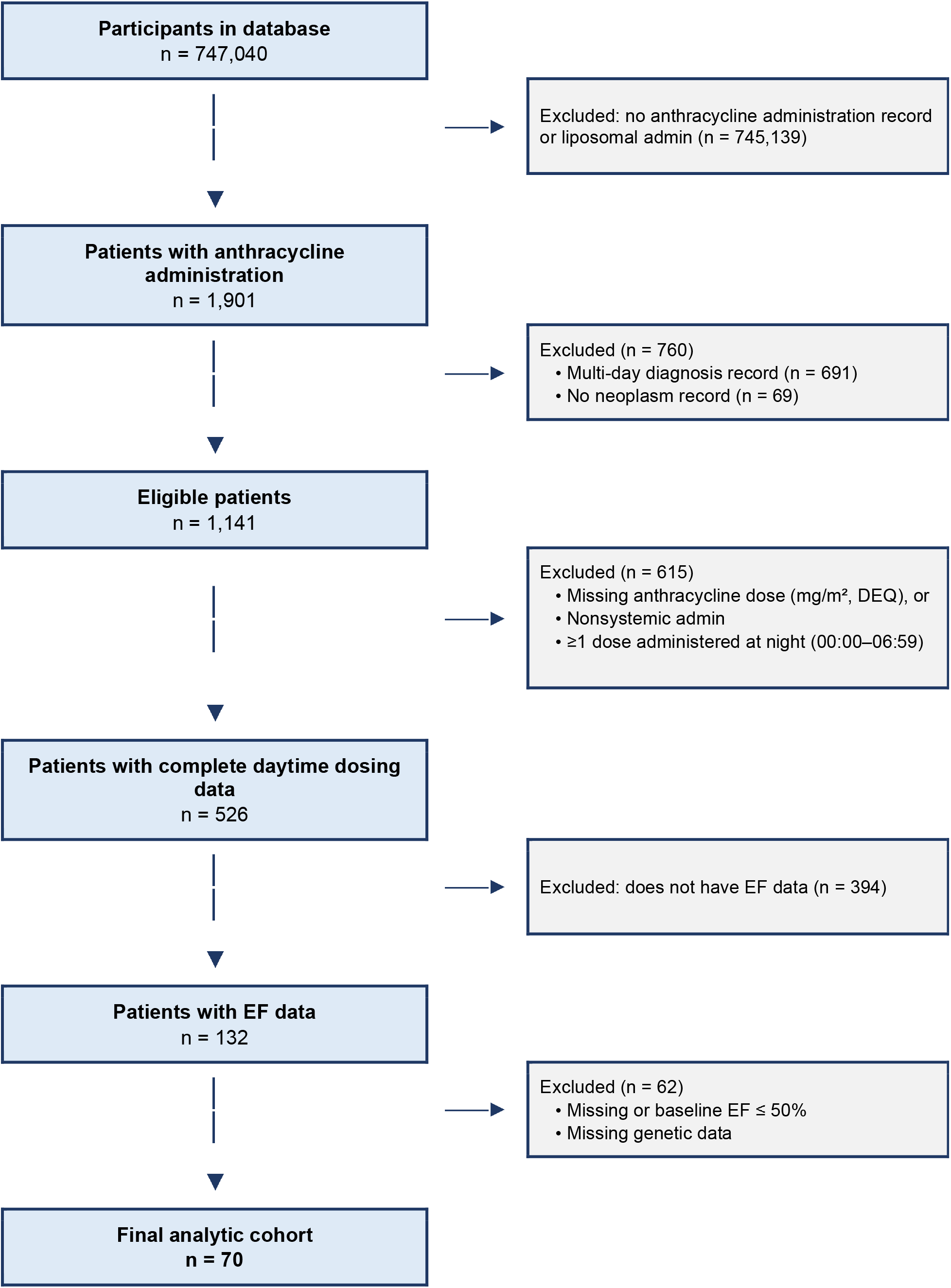
Consort style flow chart of participant selection for the All of Us (AoU) data.

**Figure S2.**
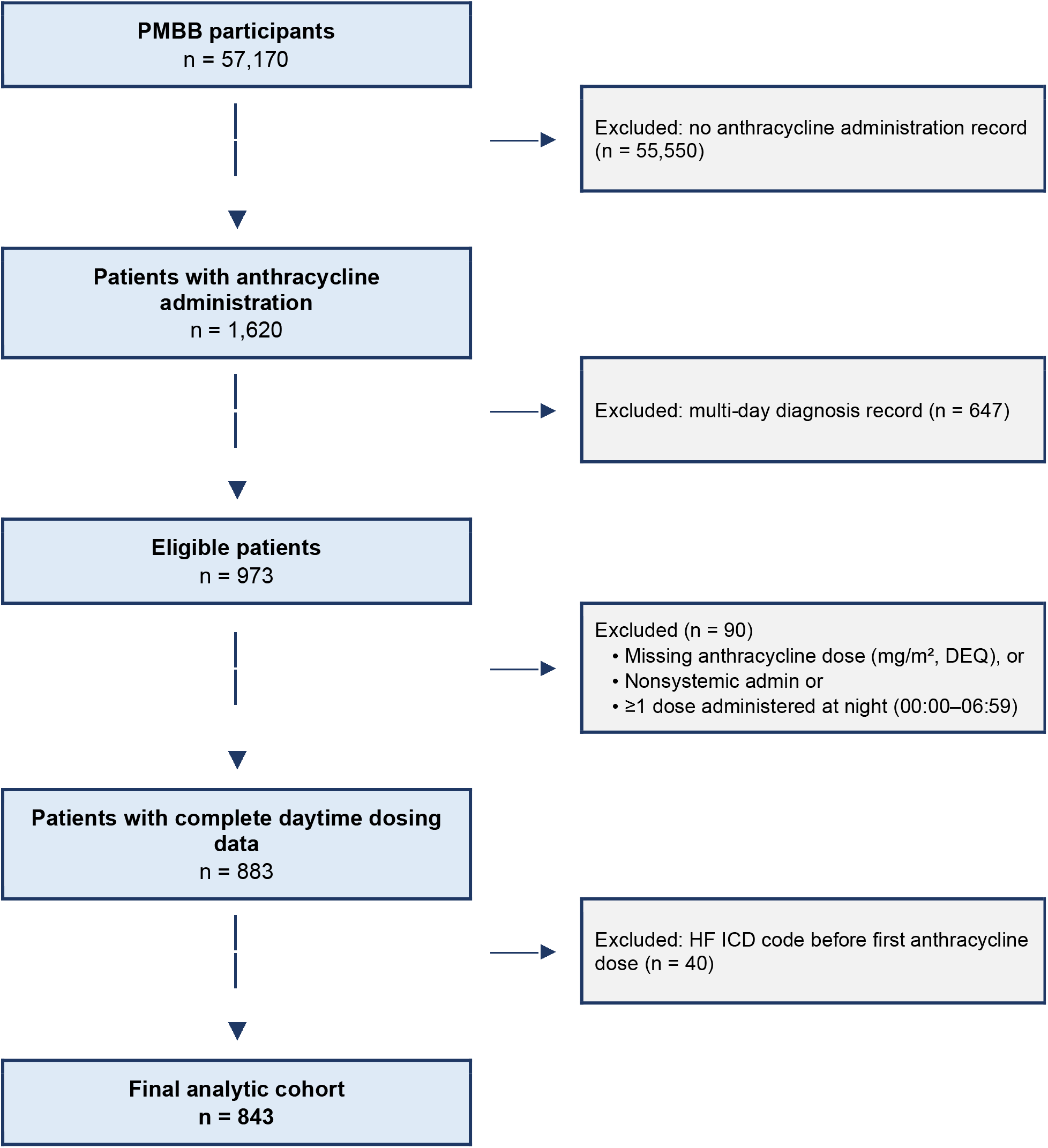
Consort style flow chart of participant selection for the Penn Medicine Biobank (PMBB) data.

**Figure S3.**
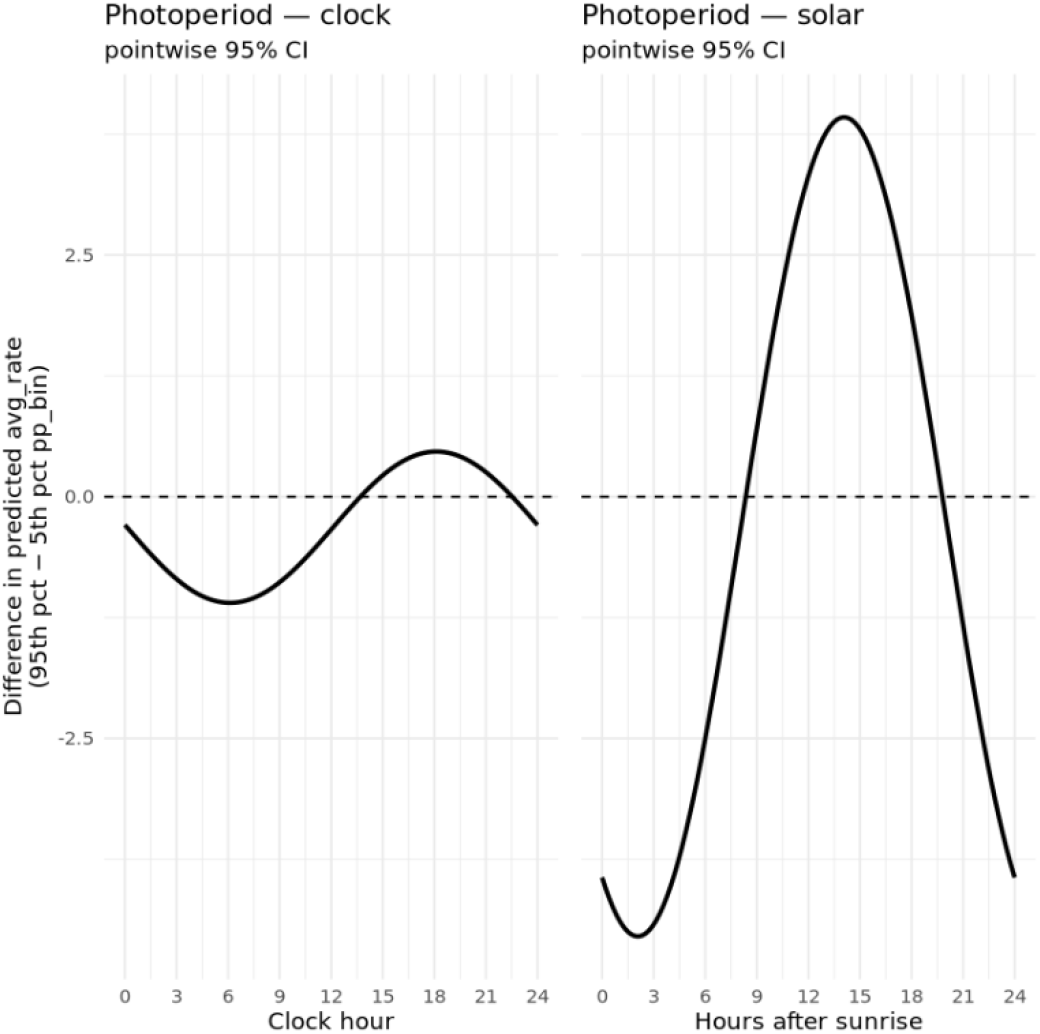
Photoperiod modification of the heart-rate rhythm, high-vs-low contrast. Difference in predicted Fitbit heart rate between the 95th and 5th photoperiod percentiles across time-of-day, with pointwise 95% confidence intervals, on the clock-time and time-from-sunrise axes (effect modification LRT p < 2e-16). Note Confidence interval ribbon is not visible due to small standard errors (mean se = 0.009435).

**Figure S4.**
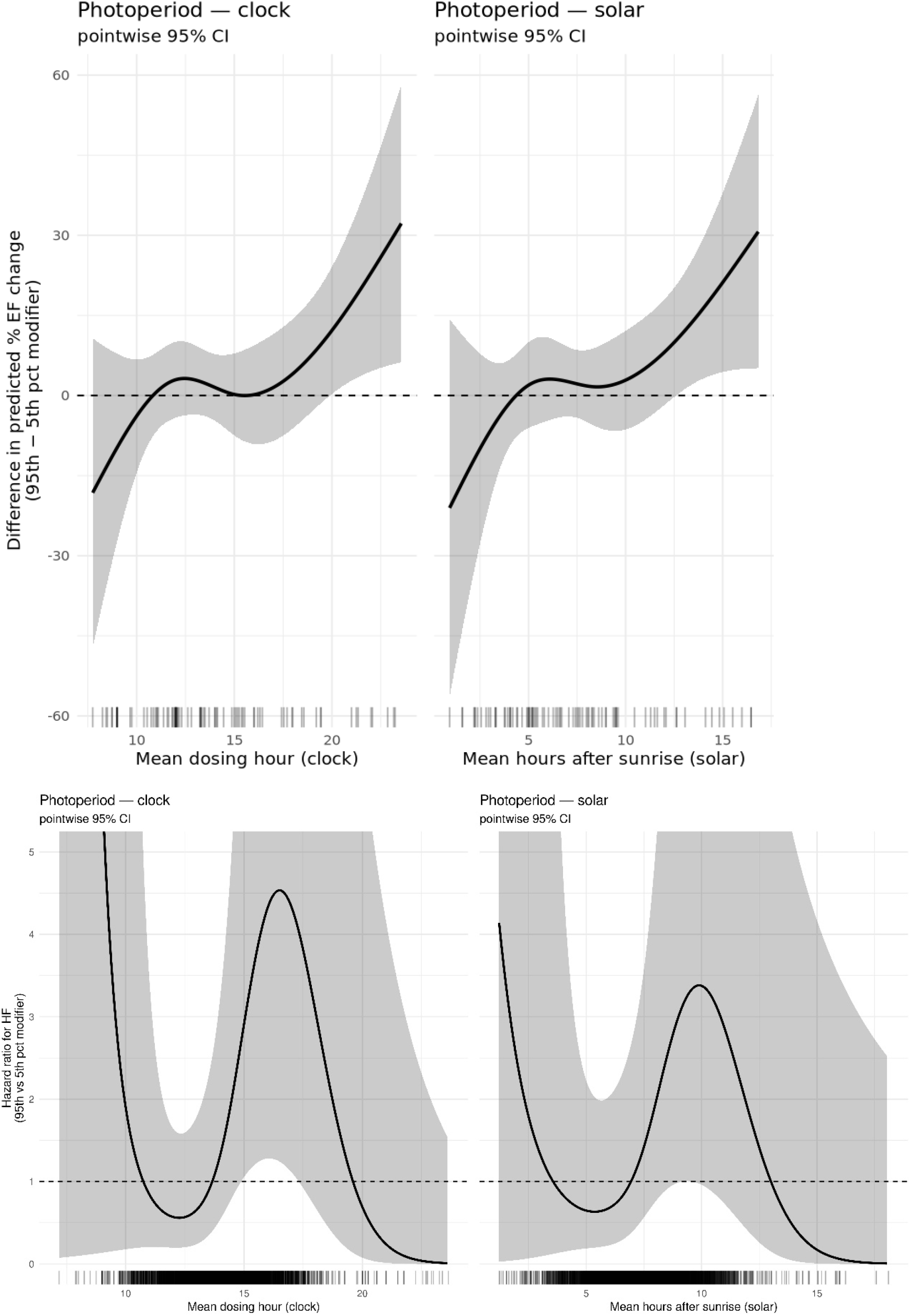
Photoperiod modification of dosing-time cardiotoxicity, high-vs-low contrast. **Top:** AoU EF Data. **Bottom:** PMBB HF data Difference in predicted outcome between the 95th and 5th photoperiod percentiles across dosing time (HF hazard ratio, PMBB, interaction LRT p = 0.023 clock, p = 0.095 sunrise; percent EF change, AoU, p = 0.064 clock, p = 0.062 sunrise), with pointwise 95% confidence intervals and per-patient rugs, shown over the full observed range of dosing times used by the likelihood-ratio tests.

**Figure S5.**
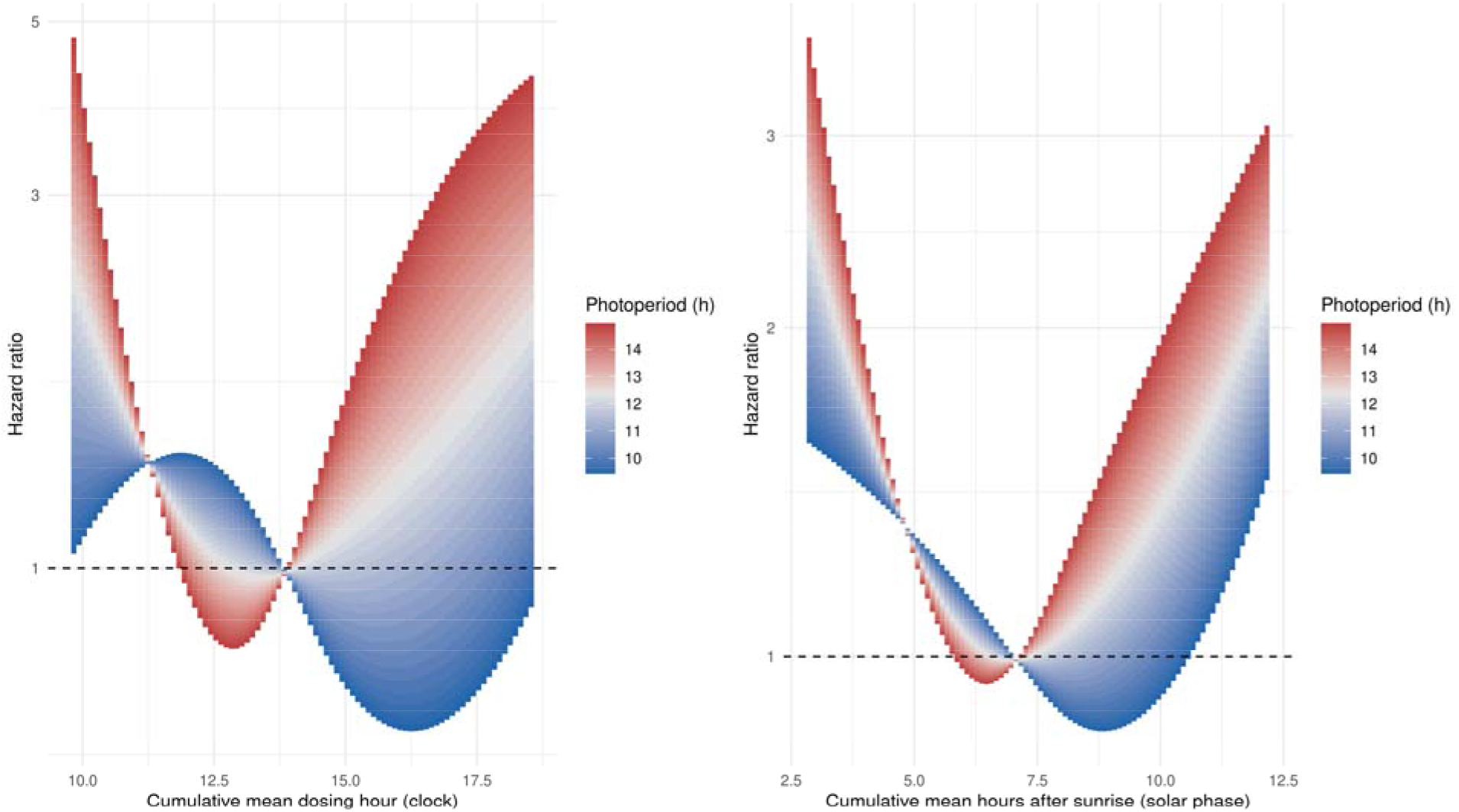
Photoperiod effect modification heat bands in the more stringent definition of HF. Hf definition consists of ICD code for HF and the prescription of a HF specific drug within 7 days (47 events). Continuous heat bands showing HF hazard ratio (PMBB; interaction LRT p = 0.414 clock, p = 0.889 sunrise) across dosing time (x-axis) and photoperiod (color scale, 5th–95th percentile), on clock-time and time-from-sunrise axes, referenced and truncated as in Figure 4. Within the data-dense interior, longer and shorter photoperiods fall on opposite sides of their respective nulls with a mid-afternoon crossover, concordant with the results with the less stringent HF definition in Figure 4.

**Figure S6.**
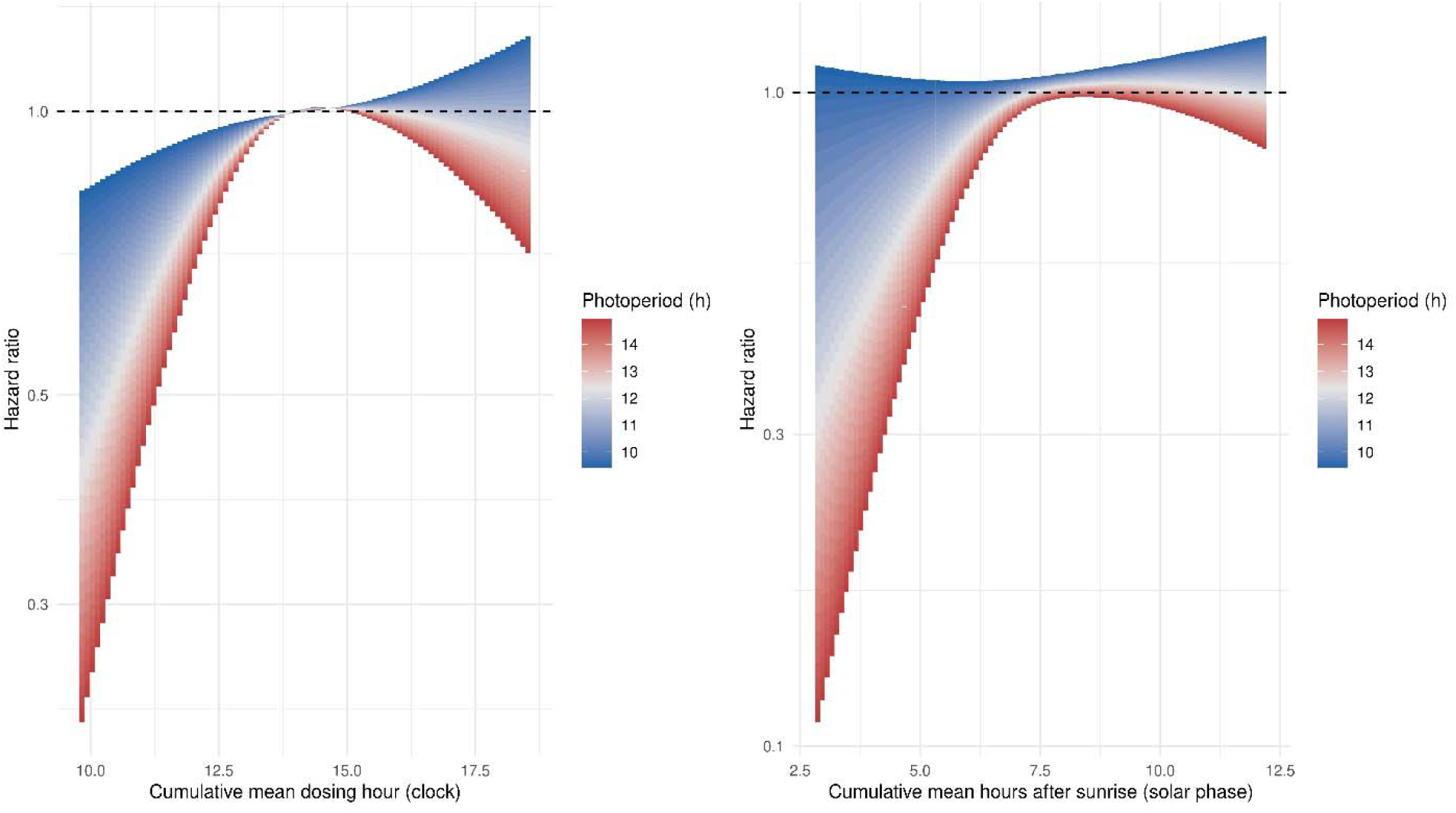
Photoperiod does not significantly modify the time-of-day effect on non-HF death. Heat bands from Cox proportional hazards models on 843 patients and 98 death events in PMBB patients without HF. Both timing axes show non-significant associations (Interaction LRT p = 0.841 for clock time and 0.789 for sunrise time).

**Figure S7.**
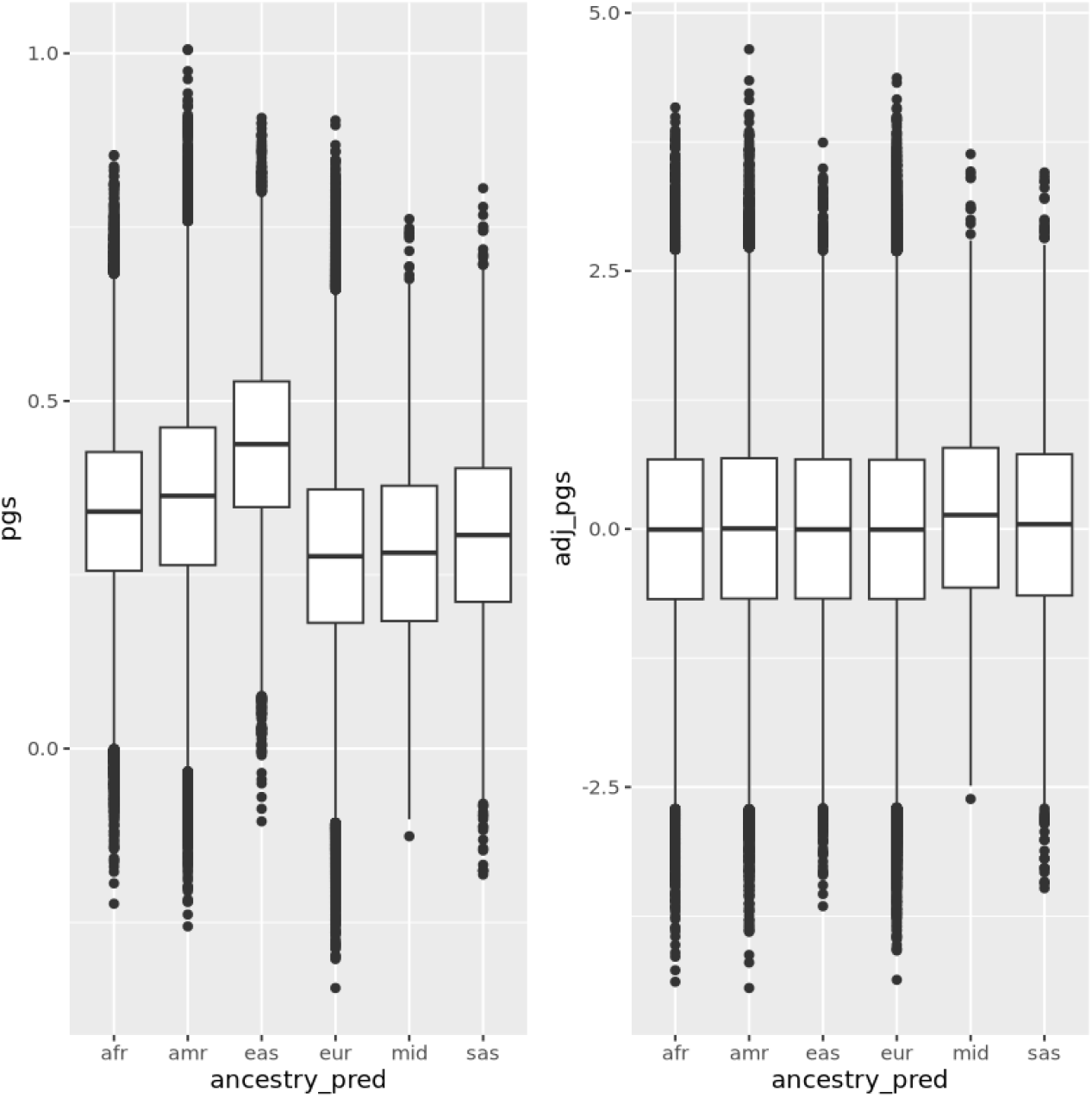
Ancestry adjustment of the chronotype polygenic score. Distribution of the raw and ancestry-adjusted PGS against predicted genetic ancestry in AoU, illustrating removal of ancestry-related mean and variance structure after adjustment.

**Figure S8.**
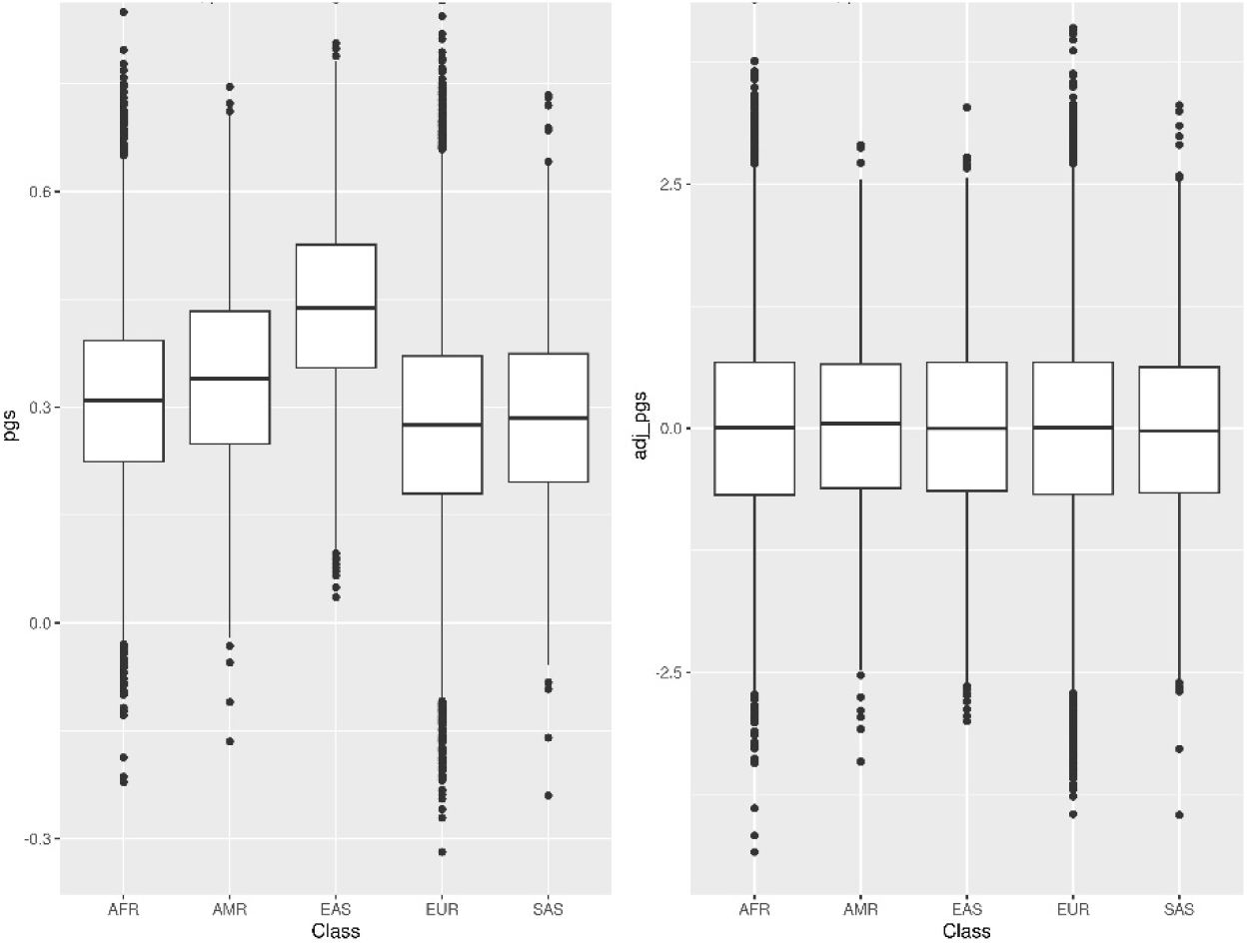
Ancestry adjustment of the chronotype polygenic score. Distribution of the raw and ancestry-adjusted PGS against predicted genetic ancestry in PMBB, illustrating removal of ancestry-related mean and variance structure after adjustment.

**Figure S9.**
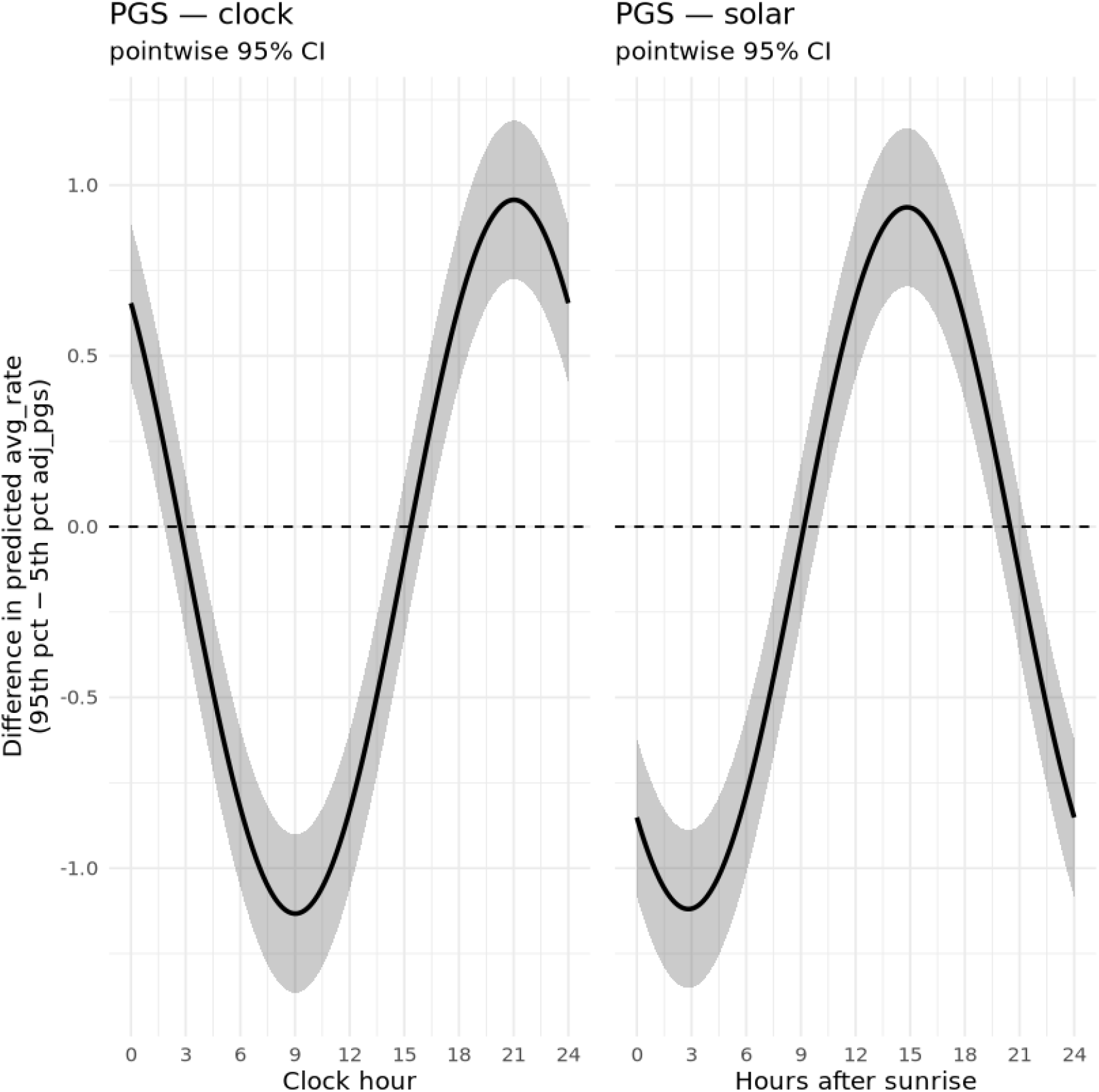
Chronotype PGS effect on the heart-rate rhythm, high-vs-low contrast. Difference in predicted Fitbit heart rate between the 95th and 5th PGS percentiles across time-of-day, with pointwise 95% confidence intervals, on the (**Left**) clock-time and (**Right**) time-from-sunrise axes (effect modification LRT p < 2e-16).

**Figure S10.**
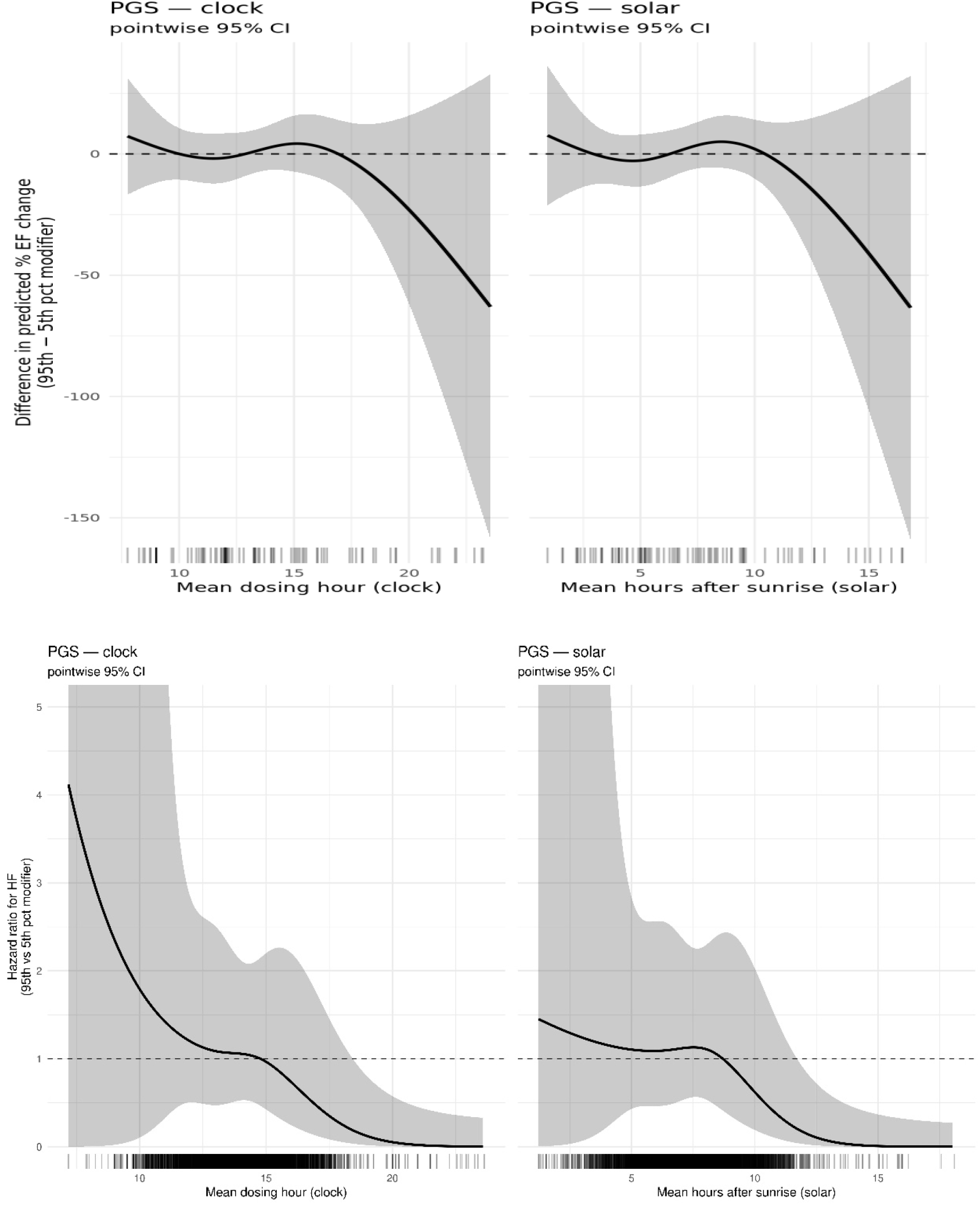
Chronotype PGS modification of dosing-time cardiotoxicity, high-vs-low contrast. **Top:** AoU EF Data. **Bottom:** PMBB HF data. Difference in predicted outcome between the 95th and 5th PGS percentiles across dosing time (HF hazard ratio, PMBB, interaction LRT p = 0.15 clock, p = 0.17 sunrise; percent EF change, AoU, p = 0.020 sunrise, p = 0.056 clock), with pointwise 95% confidence intervals and per-patient rugs, shown over the full observed range of dosing times. Wide confidence intervals and physiologically implausible point estimates at the tails indicate sparse-data extrapolation.

**Figure S11.**
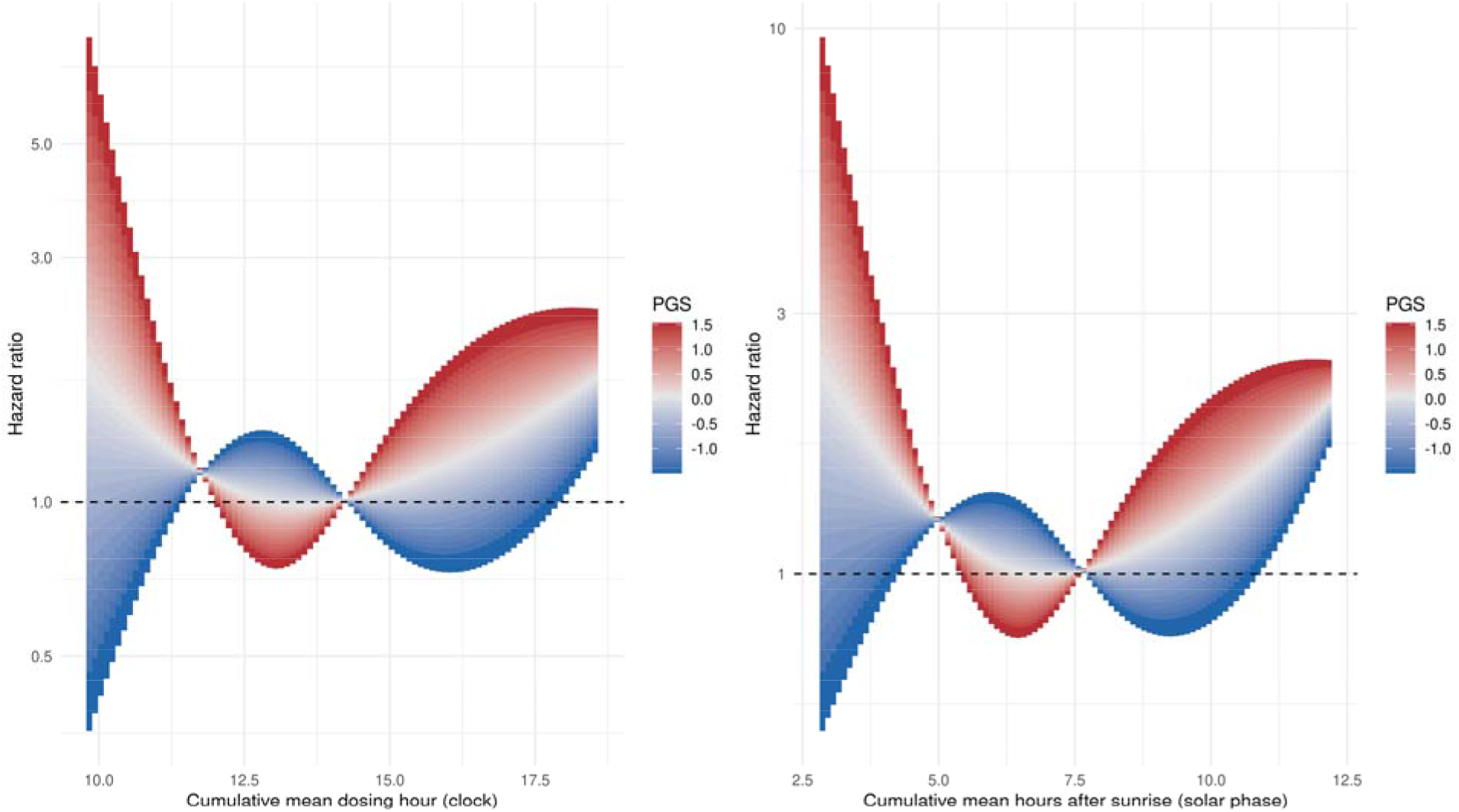
Chronotype PGS effect modification heat bands in the more stringent definition of HF. Hf definition consists of ICD code for HF and the prescription of a HF specific drug within 7 days (47 events). Continuous heat bands showing HF hazard ratio (PMBB; interaction LRT p = 0.303 clock, p = 0.345 sunrise) across dosing time (x-axis) and Chronotype PGS (color scale, 5th–95th percentile), on clock-time and time-from-sunrise axes, referenced and truncated as in Figure 4. Within the data-dense interior, longer and shorter photoperiods fall on opposite sides of their respective nulls with a mid-afternoon crossover, concordant with the results with the less stringent HF definition in Figure 6.

**Figure S12.**
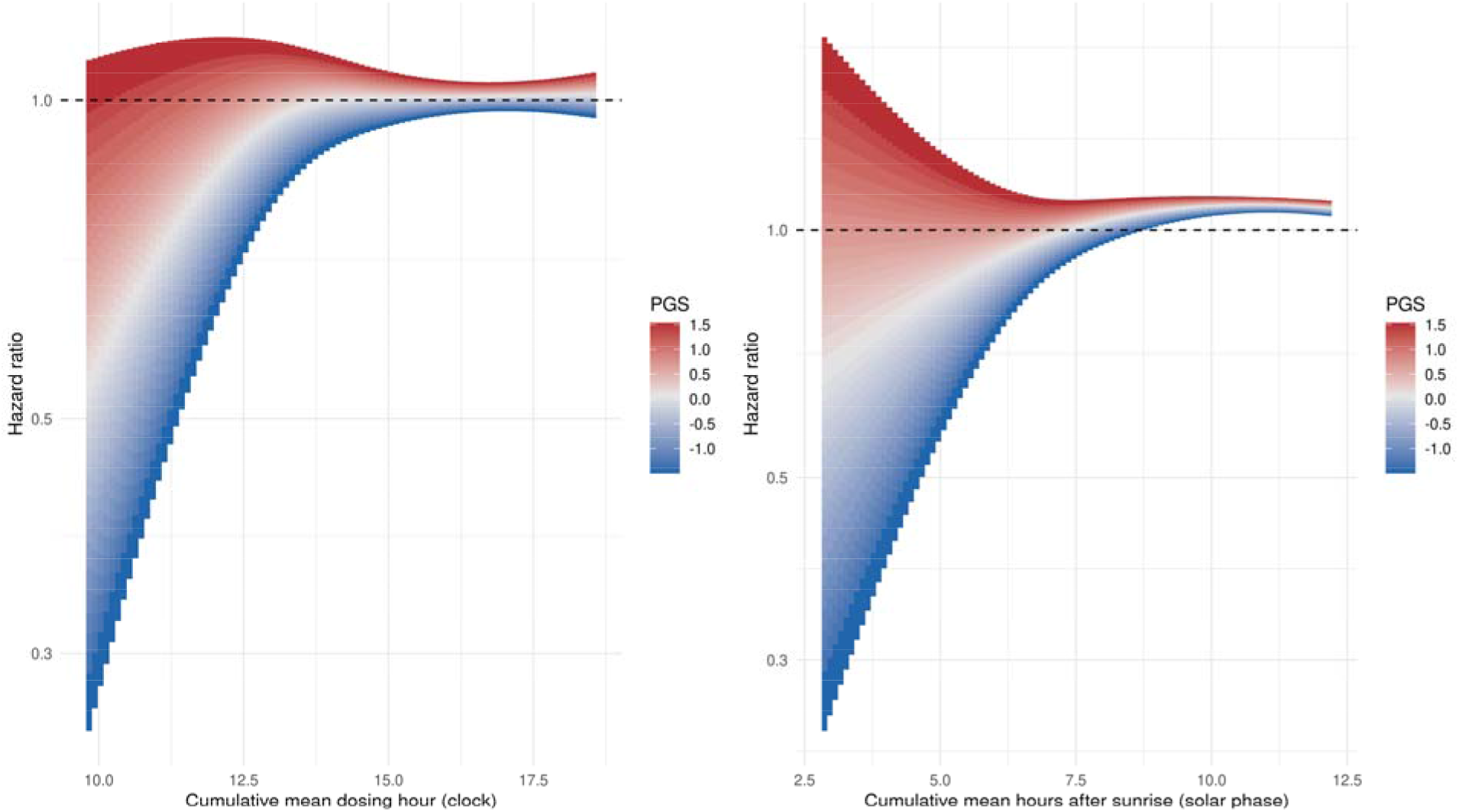
Chronotype PGS does not significantly modify the time-of-day effect on non-HF death. Heat bands from Cox proportional hazards models on 843 PMBB patients and 98 death events in patients without HF. Both timing axes show non-significant associations (Interaction LRT p = 0.918 for clock time and 0.852 for sunrise time).

## Supplementary Tables

**Table S1.** Baseline Characteristics of All of Us Ejection Fraction Analysis Cohort.

|  | level | Overall |
|---|---|---|
| n |  | 70 |
| Mean Dose Hour (median [IQR]) |  | 12.25 [10.52, 14.31] |
| Mean Time From Sunrise (median [IQR]) |  | 5.71 [3.75, 8.00] |
| Daylight Saving’s Time (%) | 5.71 [3.75, 8.00] | 284 (33.7) |
|  | TRUE | 559 (66.3) |
| Total Cumulative Dose (Mean (SD)) |  | 59.01 (24.26) |
| Age At First Dose (mean (SD)) |  | 51.44 (14.86) |
| Sex(%) | F | <20 (<14%) |
|  | M | >50 (>56%) |
| Number of echos (mean SD) |  | 2.96 (2.67) |
| Baseline EF |  | 63.62 (4.63) |
| Time of nadir echo (median [IQR] |  | 294.14 [121.74, 919.78] |
| PC1 (mean (SD)) |  | 0.07 (0.09) |
| PC2 (mean (SD)) |  | 0.07 (0.09) |
| PC3 (mean (SD)) |  | -0.02 (0.07) |

**Table S2.** Baseline Characteristics of the Penn Medicine Biobank Heart Failure Analysis Cohort.

|  | level | Overall |
|---|---|---|
| n |  | 843 |
| Mean Dose Hour (median [IQR]) |  | 13.88 [12.78, 14.89] |
| Mean Time From Sunrise (median [IQR]) |  | 7.26 [6.11, 8.43] |
| Daylight Saving’s Time (%) | FALSE | 284 (33.7) |
|  | TRUE | 559 (66.3) |
| Total Cumulative Dose (Mean (SD)) |  | 244 (116) |
| Age At First Dose (mean (SD)) |  | 48.70 (12.98) |
| Sex(%) | Female | 712 (84.5) |
|  | Male | 131 (15.5) |
| EHR encounter days(median [IQR]) |  | 245.00 [153.00, 385.50] |
| PC1 (mean (SD)) |  | 0.02 (1.01) |
| PC2 (mean (SD)) |  | 0.00 (1.05) |
| PC3 (mean (SD)) |  | 0.00 (1.01) |
| Follow up Time (median [IQR]) |  | 8.52 [4.46, 11.55] |

## Notes

### Competing Interest Statement

The authors have declared no competing interest.

### Author Declarations

All of Us research program procedures are approved by the All of US IRB and patients provide informed consent before participation. The All of Us Research Program from the National Institutes of Health determined that secondary research using All of Us Controlled Tier Data does not constitute human subject research and does not require IRB review. This study used Controlled Tier data through authorized access to the All of Us Researcher Workbench.The Penn Medicine Biobank is approved by the IRB at the University of Pennsylvania (protocol# 813913) and patients provide informed consent before participation.Our analysis of the PMBB data was deemed exempt from IRB review by the IRB at the University of Pennsylvania (protocol# 859892). For both datasets, only de-identified data was accessed and analyzed.

